# Subjective cognition trajectories, Alzheimer biomarkers, and incident mild cognitive impairment

**DOI:** 10.64898/2026.01.27.26344715

**Authors:** Elizabeth Kuhn, Luca Kleinedam, Melina Stark, Oliver Peters, Julian Hellmann-Regen, Lukas Preis, Daria Gref, Josef Priller, Eike Jakob Spruth, Maria Gemenetzi, Anja Schneider, Klaus Fliessbach, Jens Wiltfang, Claudia Bartels, Niels Hansen, Ayda Rostamzadeh, Emrah Düzel, Wenzel Glanz, Enise Incesoy, Katharina Buerger, Daniel Janowitz, Sophia Stöcklein, Robert Perneczky, Boris-Stephan Rauchmann, Stefan J. Teipel, Ingo Kilimann, Christoph Laske, Sebastian Sodenkamp, Annika Spottke, Marie Kronmüller, Sandra Roeske, Frederic Brosseron, Aflredo Ramirez, Matthis Synofzik, Matthias C. Schmid, Frank Jessen, Michael Wagner, the Alzheimer’s Disease Neuroimaging Initiative, the DELCODE study group

## Abstract

**Background:** Subjective cognitive decline (SCD) is common in older adults and may precede mild cognitive impairment (MCI). Whether longitudinal changes in self- or study partner (SP)-reported SCD improve early identification of individuals at risk for clinical progression, particularly along the Alzheimer’s disease (AD) biological continuum, remains unclear.

**Methods:** We pooled data from two longitudinal observational cohorts (DELCODE and ADNI). Cognitively unimpaired (CU) participants were recruited through public advertisement or memory clinics and included if baseline amyloid status, ≥2 SCD assessments, and clinical follow-up were available. SCD was assessed using the Everyday Cognition questionnaire (self- and SP-report). Linear mixed-effects models examined longitudinal associations between SCD trajectories, baseline AD biomarkers, and progression to incident MCI. Multivariable Cox proportional hazards models tested whether one-year changes in SCD predicted subsequent progression.

**Findings:** Among 770 participants (median age 69·9years [IQR 66·0–74·6]; 52·6% women; median follow-up 5·0years [4·0-7·0]), amyloid-positive individuals and those who progressed to MCI showed steeper longitudinal increases in both SCD reports. In amyloid-positive participants, only increases in SP-reported SCD differentiated progressors from non-progressors. One-year increases in SP-reported SCD predicted a higher risk of subsequent MCI compared with unchanged scores (hazard ratio 3·24 [95%CI 1·73-6·07]), with effects confined to amyloid-positive participants.

**Interpretation:** Longitudinal increases in SP-reported cognitive difficulties, particularly over short intervals, are associated with near-term progression to MCI in amyloid-positive CU older adults. SP-based longitudinal monitoring may represent a low-burden approach to support earlier clinical surveillance in aging populations.

**Funding:** German Center for Neurodegenerative Diseases, US National Institutes of Health.

## 1. Introduction

Subjective cognitive decline (SCD) refers to an individual’s perception of worsening cognition, despite normal performance on standardized cognitive testing.^1^ SCD is reported by nearly one-third of adults over 65,^1,2^ yet its prognostic significance remains complex. While many individuals remain cognitively stable, a substantial subset experience objective cognitive decline^3-5^ and progress to mild cognitive impairment (MCI) or dementia.^6,7^ This clinical heterogeneity underscores the need to better distinguish normal aging from early pathological trajectories.

The SCD-Initiative has identified several “SCD-plus” features associated with an increased risk for clinical progression, including the confirmation of decline by a close relative (or study partner [SP]), and, more recently, the persistence of SCD symptoms over time.^1,8,9^ Both self- and SP-reports of SCD independently predict cognitive decline, with combined reports offering greater prognostic value.^10-12^ However, as cognitive impairment advances, individuals often lose insight into their cognitive symptoms,^13,14^ rendering SP-reports increasingly important in later disease stages.

This evolving dynamic raises a key question: can longitudinal changes in self- and SP-reported SCD improve early risk prediction in cognitively unimpaired (CU) older adults compared to single-timepoint assessments? Most prior studies have relied on binary, retrospective measures of persistence (e.g., yes/no responses), which may overlook subtle or gradual changes.^15,16^ In contrast, quantitively tracking changes in symptom severity over time could provide more sensitive and dynamic indicators of risk.

Preliminary findings from the SCIENCe cohort suggest that both self- and SP-reports increased in individuals who progressed to MCI or dementia,^17^ with SP-reports increasing specifically among those with abnormal amyloid-beta (Aβ) levels, a hallmark biomarker of the Alzheimer’s disease (AD) biological continuum.^18,19^ These findings suggest that increasing SP-SCD may be especially sensitive to underlying AD pathology,^20^ yet it remains unclear whether such patterns are specific to individuals who also experience clinical progression, and whether they offer added predictive value.

In this study, we aim to validate and extend these findings across independent cohorts. We examine whether longitudinal trajectories of self- and SP-reported SCD are associated with progression to MCI in individuals with biomarker-confirmed AD pathology (i.e., abnormal Aβ alone or in combination with tau). We also test whether short-interval changes (e.g., over one year) can enhance prediction beyond baseline assessments, thereby extending the concept of persistence toward a more dynamic characterization of SCD trajectories. These insights could refine early risk stratification, inform targeted clinical surveillance, and support timely interventions in individuals at greatest risk for neurodegeneration or cognitive decline.

## 2. Methods

### 2.1. Study design

Data were obtained from two longitudinal observational studies: the *German Center for Neurodegenerative Diseases (DZNE) Longitudinal Cognitive Impairment and Dementia Study* (DELCODE, 10 university-based memory centers in Germany) and the *Alzheimer’s Disease Neuroimaging Initiative* (ADNI, 63 sites in North America, http://adni.loni.usc.edu, data retrieval on September 17, 2023 [ADNI2 and ADNI3 phases]). Both studies were approved by local ethics committees and institutional review boards, and participants provided written informed consent. DELCODE is registered with the German Clinical Trials Register (nb. DRKS00007966, 04/05/2015) and ADNI with http://clinicaltrials.gov (nb. NCT00106899). This report adheres to STROBE guidelines for observational cohort studies.

### 2.2. Participants

All selected participants were CU older adults (DELCODE, N=490; ADNI, N=280) enrolled between May 5, 2011, and November 10, 2021, who had (1) no evidence of objective cognitive impairment, (2) baseline mini mental state examination (MMSE) scores of 24-30, (3) functional activities questionnaire scores ≤9, (4) Aβ status available at baseline, and (5) underwent a clinical evaluation and completed self- and SP-reported SCD questionnaires at both baseline and at least one follow-up visit (see details in Sections **2.3-2.5**). Median (IQR) follow-up was 5·0 (4·0-7·0) years (DELCODE, 5.1 [4·0-6·6]; ADNI, 4·2 [3.0-7·5]), with a range of 1·0-8·5 years. Most participants were recruited via public advertisements; a subset of 279 DELCODE participants were patients with SCD recruited from memory clinics after reporting cognitive concerns to the referring physician. Detailed inclusion and exclusion criteria are detailed in **eMethods** and reported elsewhere.^21-23^

### 2.3. Subjective cognitive decline assessments

SCD was assessed using the Everyday Cognition Questionnaire (Ecog)^24^ in both cohorts, a 39-item questionnaire that asks the participant (self-report) and their close relative (SP-report) to rate the participant’s ability to perform an everyday task now compared to 10 years ago on a 4-point scale (from “no change” to “consistently worse”, “I don’t know” responses considered as missing values). Mean Ecog scores ranged from 1 to 4, with higher scores indicating higher SCD levels.

To ensure reliable composites scores, observations with more than 15% missing Ecog items (>5 items) were excluded. For the remaining observations, missing item-level data were imputed to allow calculation of total scores using multilevel multiple imputation (*mice* package, *2l*.*pan* method for longitudinal data), with values constrained to the original response range (1-4) to account for the hierarchical structure of repeated measures and preserve within-person variability, rather than relying on mean substitution.

A cognitive awareness index (CAI) was calculated as the difference between self- and SP-reports. Positive CAI scores indicated that the participant provided a higher rating of their difficulties compared to what their SP-reported, whereas negative scores indicated lower ratings, likely reflecting lower awareness.^25^ Cumulative score of SCD reports were also explored by summing self- and SP-reports.^3^

To quantify short-term changes, one-year difference score (hereafter called DC1) was derived by subtracting the baseline from the first follow-up score for each SCD measure. This measure was then weighted to account for the time interval between the two time points (i.e., range considered: 6-18 months). These continuous DC1 scores were categorized into “increased”, “decreased,” or “unchanged” (reference) subgroups based on whether intra-individual change exceeded ±5% of each measure’s total range (e.g., ±0·15 for mean Ecog; ±0·30 for CAI and cumulative scores). DC1 values were available in a subset of 353 (45·8%) participants (DELCODE, 215 [43·9%]; ADNI, 138 [49·3%]).

### 2.4. Clinical progression to incident-MCI

Clinical progression to incident-MCI was determined through consensus clinical review in DELCODE (biomarker- and genetic-blinded),^26^ and physician diagnoses in ADNI. A total of 131 (17·0%) participants progressed to MCI during follow-up (DELCODE, N=96 [19·6%]; ADNI, N=35 [12·5%]). Further details are available in **eMethods**.

### 2.5. Alzheimer’s disease biomarkers

Initial-stage biomarkers of Aβ and tau were assessed using multiple modalities. Among 770 participants, amyloid status was determined by Aβ-PET ([18F]florbetapir [N=221, 28·7%] or [18F]florbetaben [FBB, N=59, 7·7%]), or CSF-(N=274, 35·6%) or plasma-derived Aβ_42/40_ ratio (N=216, 28·1%). Core 1 tau status was also available in 710 participants, based on tau-PET ([18F]flortaucipir [FTP, N=106, 14·9%]), or CSF-(N=424, 59·7%) or plasma-derived ptau_181_ levels (N=180, 25·4%). Aβ+ or T1+ classifications (abnormally elevated Aβ and/or tau levels) followed cohort-specific published thresholds,^27-37^ except DELCODE plasma ptau_181_ dichotomized by Youden index (CSF ptau_181_ as reference). Overall, 239 (31·0%) participants were Aβ+, and 77 (10·9%) were Aβ+T1+. All procedures adhered to standardized cohort-specific protocols (see https://adni.bitbucket.io/reference/docs/ and **eMethods**).^27-31,33^

### 2.6. Statistical analyses

Baseline demographic, clinical, and cognitive differences were tested with Kruskall-Wallis and post-hoc Dunn tests for non-normally distributed continuous variables, and χ^2^ tests for categorical variables, across four subgroups: Aβ-Stable, Aβ+Stable, Aβ-iMCI and Aβ+iMCI, with up to eight years of follow-up.

To address our first objective, linear mixed-effects models (*lme4* package in R)^38^ with random intercepts and slopes for time (years from baseline) were used to model longitudinal changes in Ecog self- and SP-reports. Fixed effects included clinical progression to incident-MCI (model 1), baseline Aβ status (model 2), and their interaction (model 3). Three-way interactions (time x Aβ x clinical progression) were further examined with Bonferroni-corrected post hoc contrasts to test for slope differences within and across subgroups (*ggeffects* package in R).^39^ Model 1 was also replicated in Aβ stratified subgroups to determine specificity to AD. In these models, clinical progression to iMCI was modeled as a binary grouping variable in longitudinal analyses.

To address the second objective, multivariable Cox proportional hazards regression models (*survival* package) tested whether one-year SCD changes (categorical DC1, Section **2.3**) predicted clinical progression to incident-MCI (outcome, time to progression in years from the first follow-up censored at the last available assessment; model 4), and whether associations differed by baseline Aβ status (model 5). Participants converting before the first follow-up were excluded to focus on future clinical progression risk.

To isolate the incremental predictive value of DC1 beyond baseline SCD levels, multivariate Cox models including both terms were used. Kaplan-Meier curves were used for visualization. Additional multivariable models tested independent effects of self- and SP-reports.

Complementary analyses explored (1) combined SCD reports (CAI- and cumulative-Ecog scores) *versus* individual reports; and (2) whether SCD trajectories could be specific to Aβ+T_1_+ participants.

To maximize sample size, analyses were performed first in the combined sample and then repeated in stratified cohorts. All models were adjusted for age, sex, years of education, and cohort. Because Aβ and tau definitions and recruitment settings varied, these variables were added as covariates when relevant. Mixed models were additionally adjusted for the interaction of these covariates with time. Analyses were conducted using R 4.2.3 (R Foundation) from August 2022 to December 2024, with Bonferroni correction for multiple comparison across four SCD measures per model (α=0·0125).

## 3. Results

### 3.1. Participant Characteristics

Overall, 770 participants were included (median age [IQR]: 69·9 [66·0-74·6] years; 405 [52·6%] female). Participants were categorized as Aβ-stable (n=462, 60·0%), Aβ+stable (n=177, 23·0%), Aβ-iMCI (n=69, 9·0%), and Aβ+iMCI (n=62, 8·1%). Participants characteristics are summarized in **Table 1** and **eTables 1-2**.

**Table 1.**
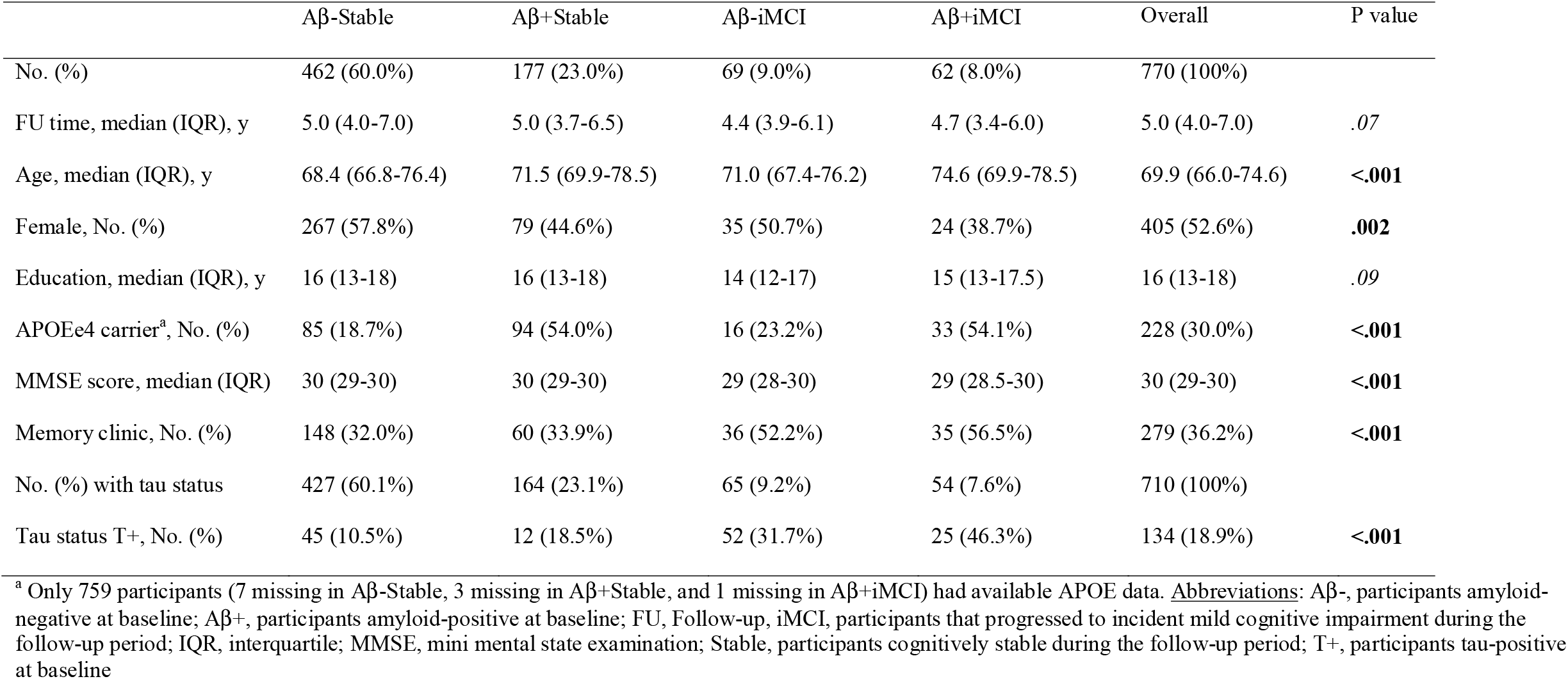
Baseline participants demographics in the combined sample (N=770) according to amyloid status and clinical progression to MCI.

Briefly, compared with Aβ-stable participants, both Aβ+ groups included fewer females and higher proportions of APOE ε4 carriers. Participants who progressed to incident MCI were more frequently recruited from memory clinics and had lower baseline MMSE scores, regardless of Aβ status. Tau positivity was more prevalent in Aβ+iMCI than in other groups, and also in Aβ+stable than in Aβ-stable participants. Education level and follow-up duration did not differ across groups.

### 3.2. Longitudinal changes in Ecog reports

At baseline, the iMCI group showed significantly higher Ecog scores than the Stable group (self-report: est.=0·08, SE=0·03, *P*=·01; SP-report: est.=0·13, SE=0·03, *P*<0·001; **Figure1A**). No baseline differences were observed by Aβ status (Self-report: est.=0·04, SE=0·03, *P*=0·17; SP-report: est.=0·009, SE=0·02, *P*=0·67; **Figure 1B**), in independent models.

**Figure 1.**
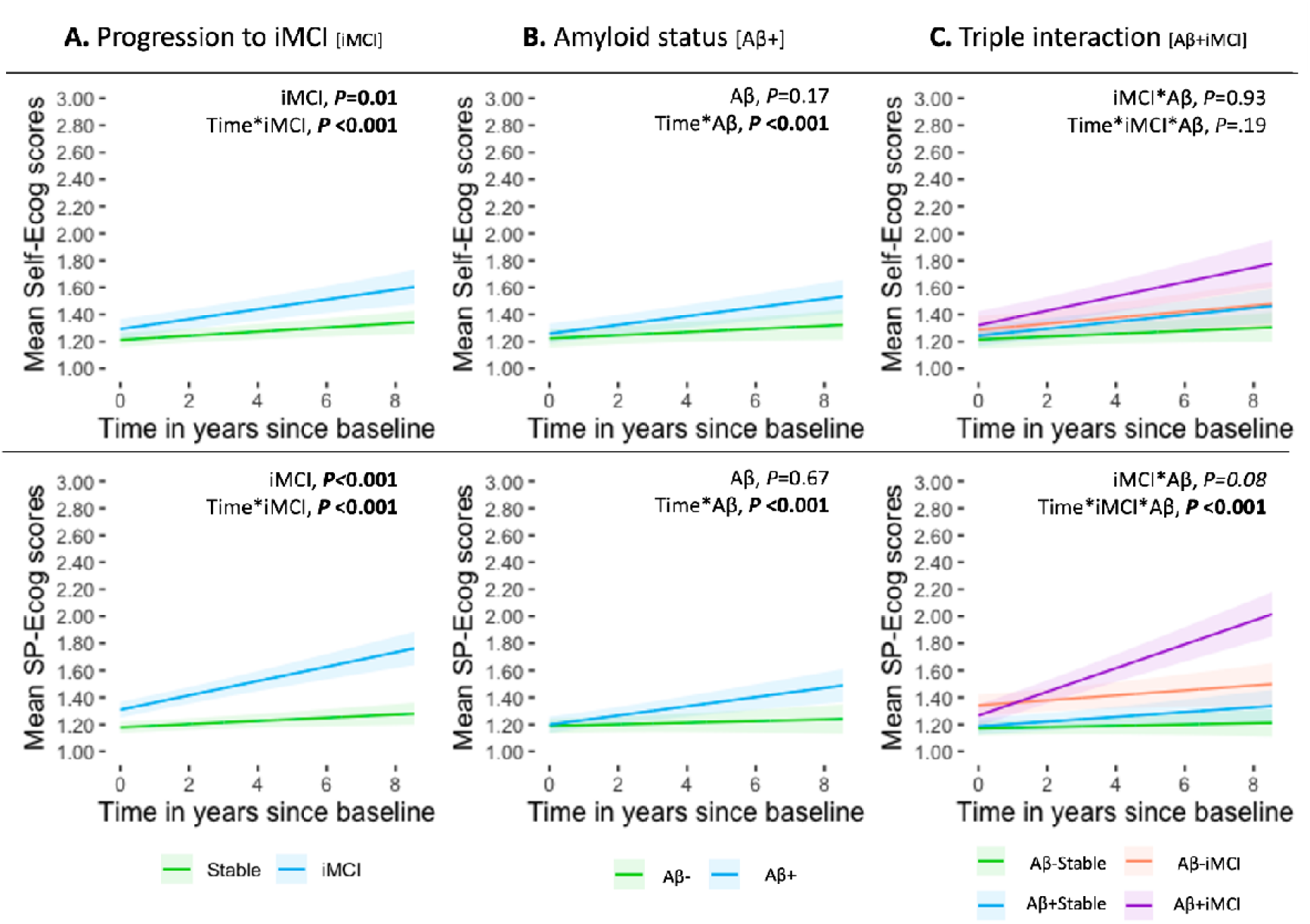
Longitudinal changes in SCD reports according to clinical progression and amyloid status. Predicted changes over time in self- and study partner (SP)-reported subjective cognitive decline (SCD) in cognitively unimpaired (CU) participants are shown according to: (A) clinical progression to mild cognitive impairment (iMCI) during the follow-up period, (B) presence of amyloid pathology (Ab+) at baseline, and (C) their interaction. The estimates illustrated here were generated using the ggeffects package of the R software.

Over time, both iMCI and Aβ+ groups demonstrated significantly steeper increases in Ecog scores compared to their respective reference groups (iMCI vs. Stable: Self-report est.=0·02, SE=0·006; SP-report est.=0·04, SE=0·006; Aβ+ vs. Aβ-: Self-report est.=0·02, SE=0·005; SP-report est.=0·03, SE=0·005; all P<0·001).

A significant three-way interaction between time, clinical progression, and Aβ status, was observed for SP-report Ecog scores only (est.=0·06, SE=0·01, P<0·001; **Figure1C**), with the steepest longitudinal increases observed in the Aβ+iMCI group (**eTable3**). Stratified analyses confirmed a significant time-by-progression interaction within the Aβ+ subgroup (est.=0·07, SE=0·01, P<0·001; **eTable4**). SP-Ecog scores increased over time in all Aβ+ subgroups but remained stable in Aβ-subgroups (**eTable3**). No significant baseline differences in Ecog scores were found among the four groups (Self-report: est.=0·006, SE=0·06, P=0·93; SP-report: est.=-0·09, SE=0·05, P=0·08). Individual trajectories are illustrated in spaghetti plots (**eFigure1)**.

Findings were broadly consistent across cohorts. In DELCODE, all main effects and interactions remained, including within the SCD subsample. In ADNI, effects surviving Bonferroni correction included the interaction between time and Aβ status on self-Ecog (est.=0·03, SE=0·009, P=0·004), and the interaction between time and clinical progression on SP-Ecog scores (baseline: est.=0·17, SE=0·05, P<0·001; slope: est.=0·06, SE=0·02, P<0·001), particularly within the Aβ+ subgroup (est.=0·07, SE=0·03, P=0·012; **eTables 3–5**).

### 3.3. Risk of progression to MCI risk according to one-year Ecog changes

DC1 SP-Ecog scores predicted future clinical progression to iMCI (P=.002), driven by increased scores (N_event_=20; HR [95% CI]=3·24[1·73-6·07]; *P*<0·001). Decreased scores showed no increased risk compared to unchanged scores (N_event_=6; HR [95% CI]=1·35[0·50-3·61]; *P*=0·55). Neither DC1 self-Ecog (*P*=0·13), nor baseline self- (HR [95% CI]=1·29[0·50-3·35]; *P*=0·59) or SP-Ecog scores (HR [95% CI]=1·11[0·43-2·85]; *P*=0·83) predicted risk (**Figure 2**).

**Figure 2.**
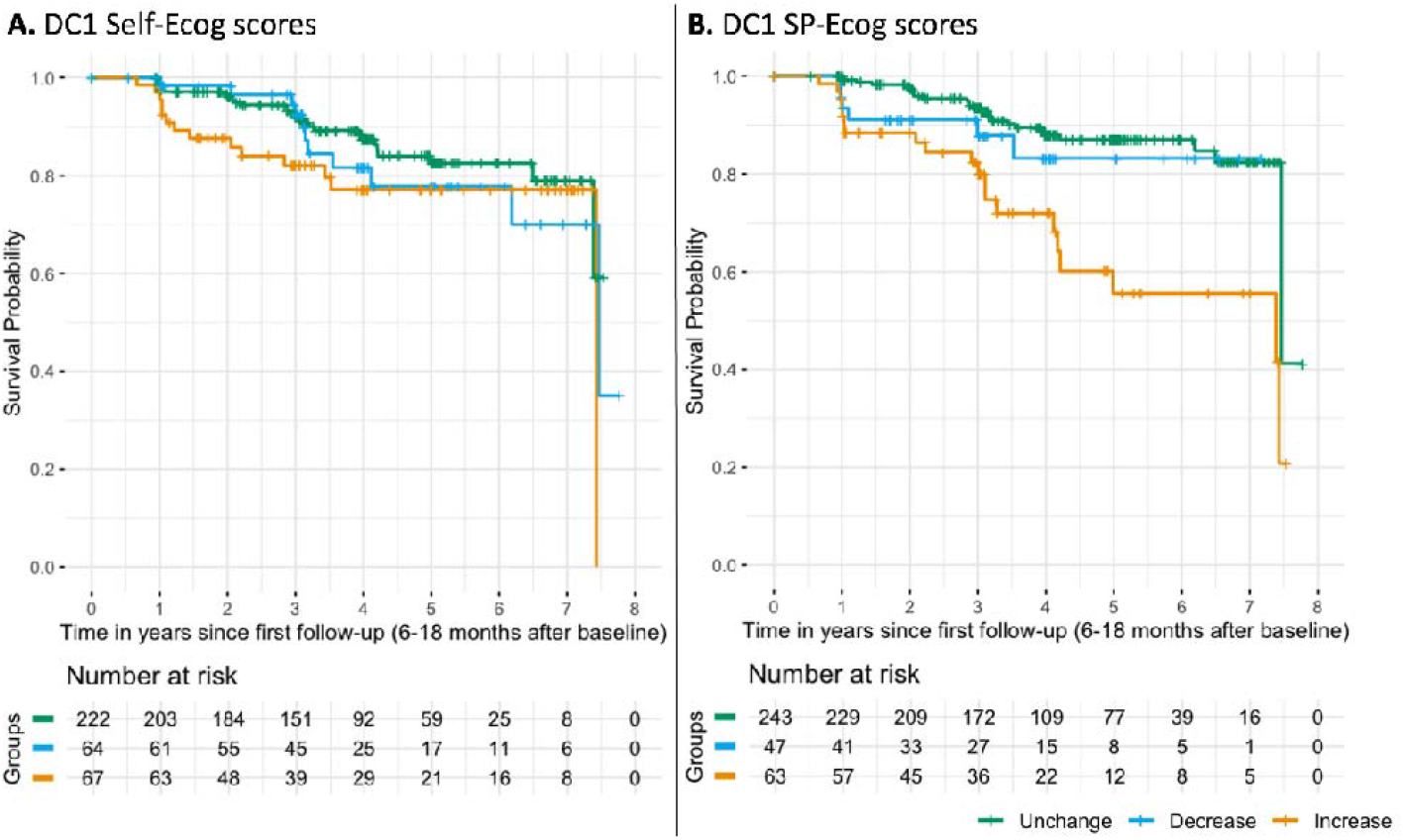
Risk of clinical progression to incident-MCI according to one-year changes in SCD reports. Kaplan-Meier curves illustrating the risk of clinical progression to incident-MCI (survival probability) from the first follow-up according to categorical DC1 scores. Groups corresponds to unchanged, decreased and increased levels of self- and study partner (SP)-Ecog scores over one-year using a 5% change threshold.

In Aβ-stratified models, increased DC1 SP-Ecog scores predicted progression in Aβ+ participants (N_event_=12; HR [95% CI]=4·18[1·80-9·73]; P<0·001), but not in Aβ-participants (*P*=0·13). However, the interaction between DC1 SP-Ecog and Aβ status was not significant (*P*=0·50). Similar trends appeared in stratified cohort analyses, although not all survived Bonferroni correction. Nothing significant was found for DC1 self-Ecog scores (**eTable6**).

In multivariate Cox models including both SP- and self-Ecog, increased DC1 SP-Ecog remained significantly associated with progression (N_event_=20; HR [95% CI]=3·93[1·82-6·49]; *P*<0·001; **eTable7**).

### 3.4. Complementary analyses

#### 3.4.1. Combined SCD reports

In linear mixed-effects models, both the iMCI and Aβ+ groups showed steeper increases in Cumulative-Ecog scores compared to Stable (est.=0·06, SE=0·009, *P*<0·001) and Aβ-(est.=0·04, SE=0·007, *P*<0·001) groups. No Bonferroni-significant effects were found for CAI-Ecog scores.

A significant three-way interaction among time, clinical progression, and Aβ status was observed, indicating a sharper decrease in CAI-Ecog scores (est.=-0·03, SE=0·01, *P*=0·009) and increase in Cumulative-Ecog scores (est.=0·07, SE=0·02, *P*<0·001) in the Aβ+iMCI group versus all others, and for Aβ+Stable versus Aβ-Stable (**Figure3A; eTable3**). Stratified analyses by Aβ status confirmed that these interactions were driven by Aβ+ participants (**eTable4**).

In Cox regression, neither DC1 Cumulative-Ecog nor CAI-Ecog scores significantly predicted progression to MCI (*P*=0·02 and *P*=0·59, respectively; **Figure 3B**; **eTable6**).

**Figure 3.**
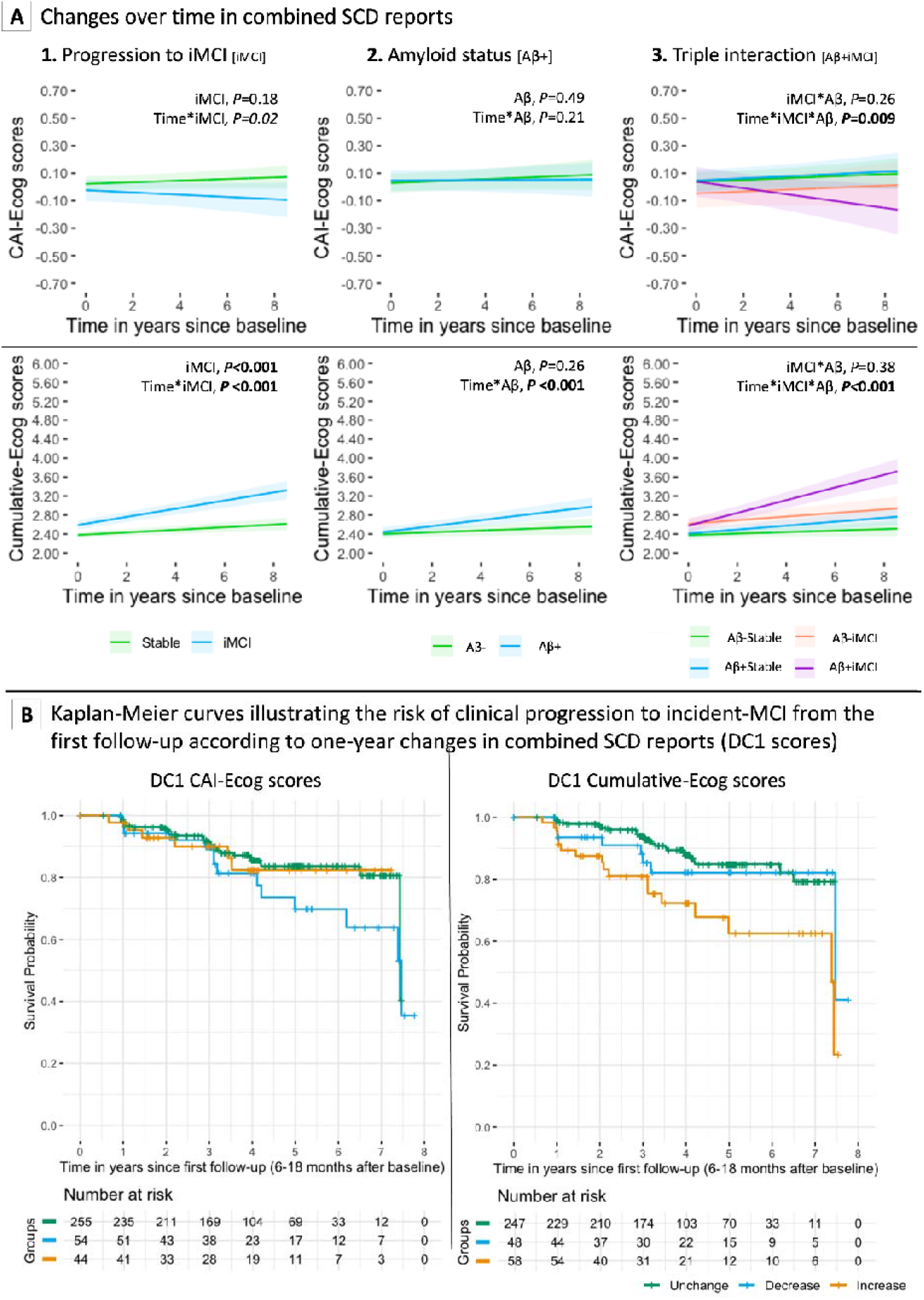
Longitudinal changes in combined SCD reports and risk of clinical progression to MCI. (A) Predicted longitudinal changes in combined subjective cognitive decline (SCD) measures among cognitively unimpaired (CU) individuals, stratified by (1) clinical progression to incident mild cognitive impairment (iMCI), (2) amyloid-β (Aβ) status at baseline, and (3) their interaction. Two SCD dimensions are presented: CAI-Ecog (discrepancy between SP– and self-reported Ecog scores) and cumulative-Ecog (combined SP- and self-reported Ecog scores). Models were adjusted for age, sex, education, and baseline Ecog scores. Shaded areas represent 95% confidence intervals. Estimates were derived using the ggeffectspackage in R. (B) Kaplan–Meier survival curves showing the risk of progression to iMCI based on one-year changes in combined SCD reports. Participants were categorized into “Decrease”, “Unchanged”, or “Increase” groups based on changes in CAI-Ecog (left panel) and cumulative-Ecog (right panel) scores between baseline and first follow-up (6–18 months). Risk groups were associated with subsequent time to progression to iMCI during longitudinal follow-up.

#### 3.4.2. By initial-stage AD biomarkers

Among participants with available tau data, longitudinal trajectories were examined according to baseline Aβ and tau status, using Aβ-T_1_-as the reference group. Significant interactions between time and AβT status were observed across all Ecog scores (all P<0·001), except CAI-Ecog (*P*=0·23**; Figure4A; eTable8**). Both Aβ+T_1_- and Aβ+T_1_+ groups showed steeper longitudinal increases in self- and SP-reports compared with Aβ-T_1_-.

**Figure 4.**
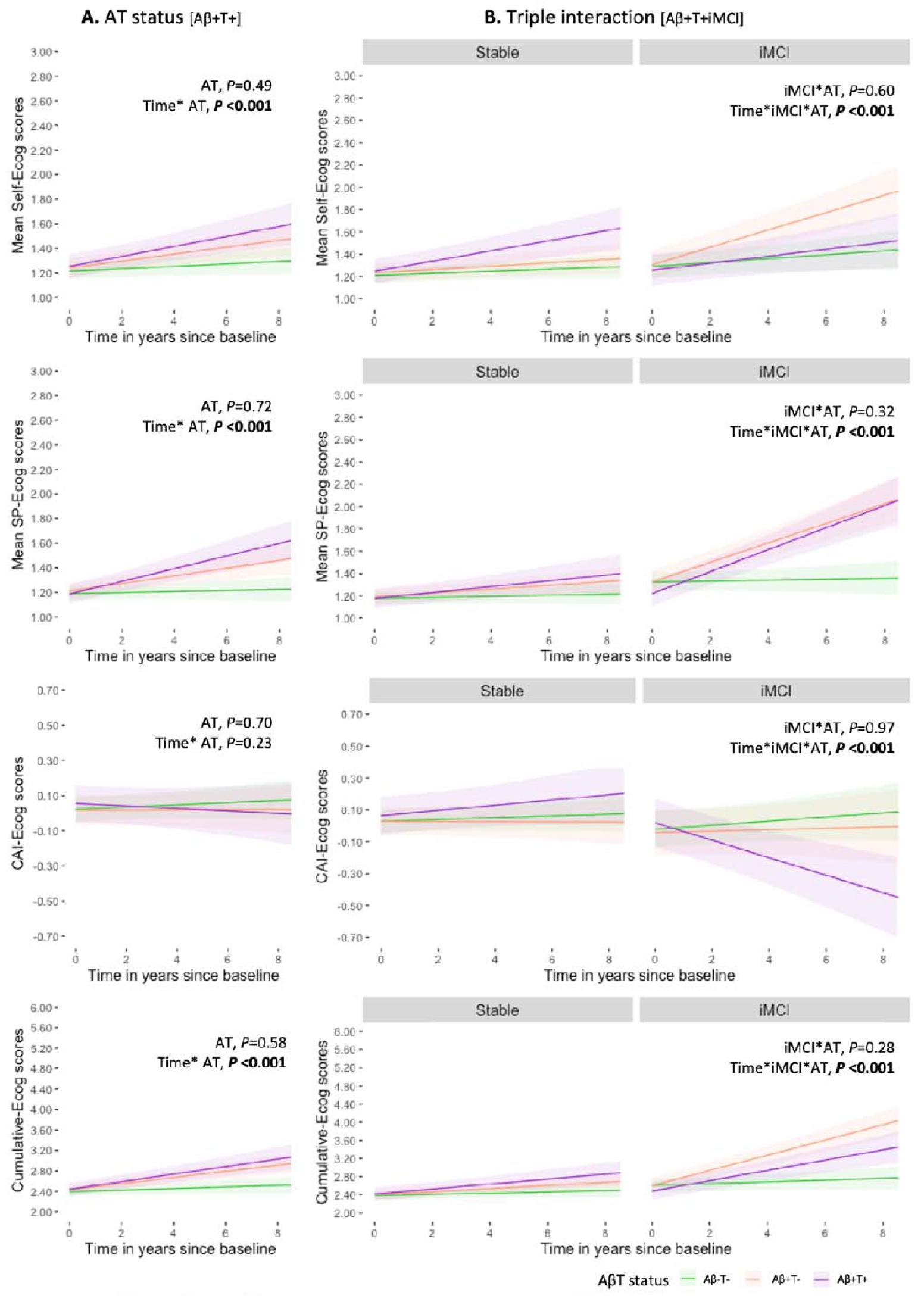
Longitudinal changes in SCD reports by clinical progression and AβT status. Predicted changes over time in self- and study partner (SP)-reported subjective cognitive decline (SCD) among cognitively unimpaired (CU) participants are shown based on baseline amyloid and tau positivity (Ab+T+; A), and its interaction with clinical progression to incident mild cognitive impairment (iMCI) during the follow-up period (B). Estimates and 95% confidence intervals were derived using the ggeffects package in R.

A significant three-way interaction (time x progression x AβT) was observed across all Ecog measures (**Figure4B**, all P<0·001). Bonferroni-adjusted post hoc comparisons indicated that both Aβ+T_1_+ and Aβ+T_1_-iMCI groups differed significantly from Stable groups and from Aβ-T_1_-iMCI on SP- and cumulative-Ecog scores (all P_adj_<0·002). For self-Ecog, differences were specific to the Aβ+T_1_-iMCI group, and for CAI-Ecog, to the Aβ+T_1_+ iMCI group (**eTable8**). Due to low event rates, Cox models stratified by AβT status were not performed.

## 4. Discussion

In this longitudinal study of CU older adults, we examined how trajectories of self- and SP-reported cognitive changes relate to baseline AD biomarkers (Aβ and tau) and subsequent progression to MCI. Across up to 8-years of follow-up, SP-reported cognitive difficulties increased more steeply in Aβ+ individuals who later progressed to MCI, whereas self-reported changes did not distinguish Aβ+ progressors from non-progressors. Importantly, similar patterns were observed over shorter time intervals: one-year increases in SP-reports predicted near-term clinical progression, with effects most evident among biomarker-positive individuals who had not yet converted during that one-year period.

At baseline, higher SCD levels (whether reported by participants themselves or by SP-) were observed among individuals who later progressed to MCI, regardless of amyloid status. In contrast, amyloid-positivity alone was not associated with greater SCD levels at study entry. Over time, however, both Aβ+ participants and clinical progressors exhibited steeper increases in self- and SP-reported SCD compared to Aβ- and clinically stable individuals. These findings align with prior studies suggesting that subtle functional-cognitive changes may be noticed by individuals and their close relatives years before formal diagnostic thresholds are reached.^11,40-42^ Beyond validating this observation, our findings may also help clarify inconsistencies in prior cross-sectional studies of SCD and Aβ status.^43-47^ They suggest a temporal dissociation whereby SCD, particularly as perceived by close relatives, may gradually intensify after amyloid reaches pathological levels and become increasingly informative and detectable as the risk of clinical progression rises. This pattern supports the notion that cross-sectional SCD severity and longitudinal change may capture distinct stages of the AD continuum.^17,48,49^ Further studies combining serial biomarker assessments with dynamic measures of cognition and awareness will be needed to clarify these temporal relationships.

A central finding of this study is that longitudinal SP-reported cognitive changes were more closely associated with clinical progression than self-reported changes in Aβ+ individuals. While both self- and SP-reports reflected increasing perception of cognitive difficulties in the presence of AD pathology, only SP-trajectories consistently differentiated those who later progressed to MCI. Informant reports are routinely used in clinical practice to characterize functional decline; our findings extend this principle by demonstrating that the rate of change in SP-reported difficulties provides prognostic information beyond baseline biomarker status, and does so well before diagnostic criteria for MCI are met. These results reinforce the added value of close relative perspectives in early disease tracking, and extend previous findings linking SP-reports to amyloid burden.^43,50,51^

Together, they suggest that longitudinal SP-based monitoring may help identify individuals at particularly high short-term risk of cognitive decline within the biologically defined AD continuum.

To further enhance clinical and public-health relevance, we examined short-term changes in perceived cognition. Previous studies have shown that persistence of self-reported SCD decline increases the likelihood of subsequent cognitive decline, but these approaches typically rely on binary indicators.^15,16^ Extending this work, we show that one-year increases in SP-reported SCD were associated with a three-to four-fold increased risk of progression to MCI among Aβ+ individuals, whereas no such associations were observed in the Aβ-group or with self-report. These findings, consistent with recent evidence from the SCIENCe cohort,^17^ support short-term SP-based change as a dynamic “SCD-plus” feature and highlight its potential value for early risk stratification in aging populations.^1,8,952^

We also explored composite metrics integrating self- and SP-reports that have been used in previous studies.^3,25^ While the Cumulative-Ecog score largely mirrored SP-report trajectories and added limited predictive value, increasing divergence between self- and SP-report (as captured by the CAI-Ecog) emerged over time among Aβ+T1+ participants who progressed to MCI. This pattern, driven by SP-reports increasingly outweighing self-perceptions, may reflect early loss of insight into cognitive difficulties accompanying AD-related neurodegeneration. However, short-term changes in CAI-Ecog did not predict clinical progression, suggesting that awareness-related measures may be more informative for characterizing disease stage than for near-term risk prediction.

Strengths of this study include the use of two large, well-characterized multicenter cohorts (DELCODE and ADNI), prospective clinical follow-up, and biomarker-confirmed AD risk stratification. The combination of long-term trajectory modeling with short-term change analyses allowed us to capture both gradual and more proximal signals of risk. However, several limitations should be acknowledged. The sample was highly educated and predominantly of European ancestry, limiting generalizability. Short-term analyses were based on smaller samples and lacked tau data and dementia conversions due to limited availability. In addition, AD biomarkers were available only at baseline, precluding evaluation of how evolving AD pathology relates to SCD trajectories. In conclusion, longitudinal increases in SP-reported SCD, particularly over short intervals, are associated with subsequent progression to MCI among amyloid-positive CU older adults. From a healthy aging perspective, these findings highlight the potential relevance of SP-based longitudinal monitoring as a low-burden approach to identifying individuals at increased short-term risk of cognitive decline before clinically meaningful impairment emerges. While further studies are needed to confirm these findings across diverse populations and settings, incorporating short-term SP-reported changes into early detection frameworks may help support more timely clinical surveillance and targeted preventive strategies aimed at delaying clinically meaningful cognitive decline in aging populations.

## Supporting information

Supplementary Online Content

## Data Availability

Data from the Alzheimer's Disease Neuroimaging Initiative (ADNI) are openly available to qualified researchers affiliated with a scientific or educational institution upon submission of a research proposal to the ADNI Data Sharing and Publications Committee (https://adni.loni.usc.edu/data-samples/adni-data/#AccessData).
Access requests for the DELCODE data can be submitted by qualified researchers to the DELCODE Steering Committee (https://www.dzne.de/en/research/research-areas/clinical-research/for-researchers/).
The analytic R code used for data processing and statistical analyses is available from the corresponding author upon reasonable request.

## Abbreviations

Aβ: Amyloid beta deposition
Aβ-: amyloid-negative
Aβ+: amyloid-positive
AD: Alzheimer’s disease
ADNI: Alzheimer’s Disease Neuroimaging Initiative
APOE: apolipoprotein E
CSF: cerebrospinal fluid
CU: cognitively unimpaired
DELCODE: DZNE Longitudinal Cognitive Impairment and Dementia Study
FBP: 18-F-Florbetapir
FTP: 18-F-Flortaucipir
MCI: patients with Mild Cognitive Impairment
NA: not available
NIA-AA: National Institute on Aging and Alzheimer’s Association
PET: Positron Emission Tomography
SCD: patients with SCD recruited from memory clinics
SD: standard deviation
SUVr: global standardized uptake value ratio
T-: tau-negative
T+: tau-positive.

## Acknowledgements

^*^ Part of the data used in preparation of this article was obtained from the Alzheimer’s Disease Neuroimaging Initiative (ADNI) database (https://adni.loni.usc.edu). As such, the investigators within the ADNI contributed to the design and implementation of ADNI and/or provided data, but they did not participate in analysis or writing of this report. A complete listing of ADNI investigators can be found at: http://adni.loni.usc.edu/wp-content/uploads/how_to_apply/ADNI_Acknowledgement_List.pdf. The ADNI was launched in 2003 as a public-private partnership led by Principal Investigator Michael W. Weiner, MD. The primary goal of ADNI has been to test whether serial magnetic resonance imaging (MRI), positron emission tomography (PET), other biological markers, and clinical and neuropsychological assessment can be combined to measure the progression of mild cognitive impairment (MCI) and early Alzheimer’s disease (AD).

^§^ Part of data used in the preparation of this article were obtained from the DELCODE study (https://www.dzne.de/en/research/studies/clinical-studies/delcode/). The authors thank the following institutions: Max-Delbrück-centrum für Molekulare medizin in der Helmholtz-Gemeinschaft (MDC), Freie Universität Berlin Center for Cognitive neuroscience Berlin (CCNB), Klinik und Poliklinik für Nuklearmedizin. Universitätsklinikum Bonn. Venusberg-Campus 1, Uniklinik Köln - Klinik und Poliklinik für Nuklearmedizin, Universitätsklinik Magdeburg-Zentrum für Radiologie Klinik für Radiologie und Nuklearmedizin, Klinik und Poliklinik für Nuklearmedizin Klinikum der Universität München, Universitätsklinikum Rostock Klinik und Poliklinik für Nuklearmedizin, Nuklearmedizin und Klinische Molekulare Bildgebung - Univeristätsklinikum Tübingen, Bernstein Center für Computational Neuroscience Berlin, Universitätsmedizin Göttingen Core Facility MR-Research Göttingen, Institut für Klinische Radiologie Klinikum der Universität München, Universitätsklinikum Tübingen MR-Forschungszentrum.

## Conflicts of interest

All the authors declare no conflict of interest.

## Funding Sources

EK was funded by the Fondation Philippe Chatrier and the Helmholtz Artificial Intelligence Cooperation Unit. MS was funded by a Hertie Network of Excellence in Clinical Neuroscience research grant awarded to LK.

^*^ Alzheimer’s Disease Neuroimaging Initiative data collection and sharing for this project was funded by the ADNI (National Institutes of Health Grant U01 AG024904) and DOD ADNI (Department of Defense award number W81XWH-12-2-0012). ADNI is funded by the National Institute on Aging, the National Institute of Biomedical Imaging and Bioengineering, and through generous contributions from the following: AbbVie, Alzheimer’s Association; Alzheimer’s Drug Discovery Foundation; Araclon Biotech; BioClinica, Inc.; Biogen; Bristol-Myers Squibb Company; CereSpir, Inc.; Cogstate; Eisai Inc.; Elan Pharmaceuticals, Inc.; Eli Lilly and Company; EuroImmun; F. Hoffmann-La Roche Ltd and its affiliated company Genentech, Inc.; Fujirebio; GE Healthcare; IXICO Ltd.; Janssen Alzheimer Immunotherapy Research & Development, LLC.; Johnson & Johnson Pharmaceutical Research & Development LLC.; Lumosity; Lundbeck; Merck & Co., Inc.; Meso Scale Diagnostics, LLC.; NeuroRx Research; Neurotrack Technologies; Novartis Pharmaceuticals Corporation; Pfizer Inc.; Piramal Imaging; Servier; Takeda Pharmaceutical Company; and Transition Therapeutics. The Canadian Institutes of Health Research is providing funds to support ADNI clinical sites in Canada. Private sector contributions are facilitated by the Foundation for the National Institutes of Health (www.fnih.org). The grantee organization is the Northern California Institute for Research and Education, and the study is coordinated by the Alzheimer’s Therapeutic Research Institute at the University of Southern California. ADNI data are disseminated by the Laboratory for Neuro Imaging at the University of Southern California.

^§^ The DELCODE study was funded by the German Center for Neurodegenerative Diseases (Deutsches Zentrum fu□r Neurodegenerative Erkrankungen (DZNE)), reference number BN012.

## Notes

### Competing Interest Statement

The authors have declared no competing interest.

### Author Declarations

Ethics committees and Institutional Review Boards of all participating institutions of the German Center for Neurodegenerative Diseases (DZNE) gave ethical approval for the DELCODE study protocol (German Clinical Trials Register [DRKS00007966]). Institutional Review Boards of all participating institutions in the Alzheimer's Disease Neuroimaging Initiative (ADNI) gave ethical approval for the ADNI study protocol (http://clinicaltrials.gov [nb. NCT00106899]). All datasets used in this study were de-identified prior to their use. No personally identifiable information was included in the data analyzed for this research.

## References

1. Jessen F, Amariglio RE, van Boxtel M, et al. A conceptual framework for research on subjective cognitive decline in preclinical Alzheimer’s disease. Alzheimer’s & Dementia: The Journal of the Alzheimer’s Association. 2014/11// 2014;10(6):844–852. doi:10.1016/j.jalz.2014.01.001

2. Röhr S, Pabst A, Riedel-Heller SG, et al. Estimating prevalence of subjective cognitive decline in and across international cohort studies of aging: a COSMIC study. Alzheimers Res Ther. 12 2020;12(1):167. doi:10.1186/s13195-020-00734-y

3. Amariglio RE, Donohue MC, Marshall GA, et al. Tracking early decline in cognitive function in older individuals at risk for Alzheimer’s disease dementia: the Alzheimer’s Disease Cooperative Study Cognitive Function Instrument. JAMA neurology. 2015/04/01/ 2015;72(4):446–454. doi:10.1001/jamaneurol.2014.3375

4. Gifford KA, Liu D, Carmona H, et al. Inclusion of an informant yields strong associations between cognitive complaint and longitudinal cognitive outcomes in non-demented elders. Journal of Alzheimer’s disease : JAD. 2015 2015;43(1):121–132. doi:10.3233/JAD-131925

5. Koppara A, Wagner M, Lange C, et al. Cognitive performance before and after the onset of subjective cognitive decline in old age. Alzheimer’s & Dementia : Diagnosis, Assessment & Disease Monitoring. 2015/05/02/ 2015;1(2):194–205. doi:10.1016/j.dadm.2015.02.005

6. Reisberg B, Shulman MB, Torossian C, Leng L, Zhu W. Outcome over seven years of healthy adults with and without subjective cognitive impairment. Alzheimer’s & dementia : the journal of the Alzheimer’s Association. 2010/01// 2010;6(1)doi:10.1016/j.jalz.2009.10.002

7. Wang XT, Wang ZT, Hu HY, et al. Association of Subjective Cognitive Decline with Risk of Cognitive Impairment and Dementia: A Systematic Review and Meta-Analysis of Prospective Longitudinal Studies. J Prev Alzheimers Dis. 2021;8(3):277–285. doi:10.14283/jpad.2021.27

8. Molinuevo JL, Rabin LA, Amariglio R, et al. Implementation of subjective cognitive decline criteria in research studies. Alzheimer’s & Dementia: The Journal of the Alzheimer’s Association. 2017/03// 2017;13(3):296–311. doi:10.1016/j.jalz.2016.09.012

9. Jessen F, Amariglio RE, Buckley RF, et al. The characterisation of subjective cognitive decline. The Lancet Neurology. 2020/03// 2020;19(3):271–278. doi:10.1016/S1474-4422(19)30368-0

10. Nosheny RL, Jin C, Neuhaus J, et al. Study partner-reported decline identifies cognitive decline and dementia risk. Ann Clin Transl Neurol. 12 2019;6(12):2448–2459. doi:10.1002/acn3.50938

11. Nosheny RL, Amariglio R, Sikkes SAM, et al. The role of dyadic cognitive report and subjective cognitive decline in early ADRD clinical research and trials: Current knowledge, gaps, and recommendations. Alzheimer’s & Dementia: Translational Research & Clinical Interventions. 2022/01/01 2022;8(1):e12357. doi:10.1002/trc2.12357

12. Caselli RJ, Chen K, Locke DEC, et al. Subjective cognitive decline: self and informant comparisons. Alzheimer’s & Dementia: The Journal of the Alzheimer’s Association. 2014/01// 2014;10(1):93–98. doi:10.1016/j.jalz.2013.01.003

13. Starkstein SE. Anosognosia in Alzheimer’s disease: Diagnosis, frequency, mechanism and clinical correlates. Cortex. 2014/12/01/ 2014;61:64-73. Special Issue: Understanding Babinski’s anosognosia: 100 years later. doi:10.1016/j.cortex.2014.07.019

14. Wilson RS, Sytsma J, Barnes LL, Boyle PA. Anosognosia in Dementia. Current Neurology and Neuroscience Reports. 2016//09/ 2016;16(9)(Article 77):1–6. doi:10.1007/s11910-016-0684-z

15. Wolfsgruber S, Kleineidam L, Wagner M, et al. Differential Risk of Incident Alzheimer’s Disease Dementia in Stable Versus Unstable Patterns of Subjective Cognitive Decline. Journal of Alzheimer’s disease: JAD. 2016//10/04 2016;54(3):1135–1146. doi:10.3233/JAD-160407

16. Roehr S, Villringer A, Angermeyer MC, Luck T, Riedel-Heller SG. Outcomes of stable and unstable patterns of subjective cognitive decline – results from the Leipzig Longitudinal Study of the Aged (LEILA75+). BMC Geriatrics. 2016/11/04/ 2016;16(1):180. doi:10.1186/s12877-016-0353-8

17. Dubbelman MA, Sikkes SAM, Ebenau JL, et al. Changes in self- and study partner-perceived cognitive functioning in relation to amyloid status and future clinical progression: Findings from the SCIENCe project. Alzheimers Dement. Jul 2023;19(7):2933–2942. doi:10.1002/alz.12931

18. Jack CR, Bennett DA, Blennow K, et al. NIA-AA Research Framework: Toward a biological definition of Alzheimer’s disease. Alzheimer’s & Dementia: The Journal of the Alzheimer’s Association. 2018//04/ 2018;14(4):535–562. doi:10.1016/j.jalz.2018.02.018

19. Jack CR, Andrews JS, Beach TG, et al. Revised criteria for diagnosis and staging of Alzheimer’s disease: Alzheimer’s Association Workgroup. Alzheimers Dement. Jun 27 2024;doi:10.1002/alz.13859

20. Kuhn E, Perrotin A, La Joie R, et al. Association of the Informant-Reported Memory Decline With Cognitive and Brain Deterioration Through the Alzheimer Clinical Continuum. Neurology. Jun 13 2023;100(24):e2454–e2465. doi:10.1212/WNL.0000000000207338

21. Jessen F, Spottke A, Boecker H, et al. Design and first baseline data of the DZNE multicenter observational study on predementia Alzheimer’s disease (DELCODE). Alzheimers Res Ther. 02 2018;10(1):15. doi:10.1186/s13195-017-0314-2

22. Petersen RC, Aisen PS, Beckett LA, et al. Alzheimer’s Disease Neuroimaging Initiative (ADNI): clinical characterization. Neurology. Jan 2010;74(3):201–9. doi:10.1212/WNL.0b013e3181cb3e25

23. Aisen PS, Petersen RC, Donohue MC, et al. Clinical core of the Alzheimer’s disease neuroimaging initiative: Progress and plans. Alzheimer’s & Dementia. 2010/05/01/ 2010;6(3):239–246. doi:10.1016/j.jalz.2010.03.006

24. Farias ST, Mungas D, Reed BR, et al. The measurement of everyday cognition (ECog): scale development and psychometric properties. Neuropsychology. Jul 2008;22(4):531–44. doi:10.1037/0894-4105.22.4.531

25. Ryu SY, Kim A, Kim S, et al. Self- and informant-reported cognitive functioning and awareness in subjective cognitive decline, mild cognitive impairment, and very mild Alzheimer disease. International Journal of Geriatric Psychiatry. 2020//01/ 2020;35(1):91–98. doi:10.1002/gps.5224

26. Stark M, Wolfsgruber S, Kleineidam L, et al. Relevance of Minor Neuropsychological Deficits in Patients With Subjective Cognitive Decline. Neurology. Nov 21 2023;101(21):e2185–e2196. doi:10.1212/WNL.0000000000207844

27. Jessen F, Wolfsgruber S, Kleineindam L, et al. Subjective cognitive decline and stage 2 of Alzheimer disease in patients from memory centers. Alzheimers Dement. Feb 2023;19(2):487–497. doi:10.1002/alz.12674

28. Vogelgsang J, Hansen N, Stark M, et al. Plasma amyloid beta X-42/X-40 ratio and cognitive decline in suspected early and preclinical Alzheimer’s disease. Alzheimers Dement. Aug 2024;20(8):5132–5142. doi:10.1002/alz.13909

29. Navitsky M, Joshi AD, Kennedy I, et al. Standardization of amyloid quantitation with florbetapir standardized uptake value ratios to the Centiloid scale. Alzheimers Dement. Dec 2018;14(12):1565–1571. doi:10.1016/j.jalz.2018.06.1353

30. Rowe CC, Doré V, Jones G, et al. F-Florbetaben PET beta-amyloid binding expressed in Centiloids. Eur J Nucl Med Mol Imaging. Nov 2017;44(12):2053–2059. doi:10.1007/s00259-017-3749-6

31. Aksman LM, Oxtoby NP, Scelsi MA, et al. A data-driven study of Alzheimer’s disease related amyloid and tau pathology progression. Brain. Dec 01 2023;146(12):4935–4948. doi:10.1093/brain/awad232

32. Bertens D, Tijms BM, Scheltens P, Teunissen CE, Visser PJ. Unbiased estimates of cerebrospinal fluid β-amyloid 1-42 cutoffs in a large memory clinic population. Alzheimers Res Ther. Feb 14 2017;9(1):8. doi:10.1186/s13195-016-0233-7

33. Mengel D, Soter E, Ott JM, et al. Blood biomarkers confirm subjective cognitive decline (SCD) as a distinct molecular and clinical stage within the NIA-AA framework of Alzheimer’s disease. medRxiv. 2024:2024.07.10.24310205. doi:10.1101/2024.07.10.24310205

34. Landau SM, Mintun MA, Joshi AD, et al. Amyloid deposition, hypometabolism, and longitudinal cognitive decline. Ann Neurol. Oct 2012;72(4):578–86. doi:10.1002/ana.23650

35. Landau SM, Lu M, Joshi AD, et al. Comparing positron emission tomography imaging and cerebrospinal fluid measurements of β-amyloid. Ann Neurol. Dec 2013;74(6):826–36. doi:10.1002/ana.23908

36. Ge X, Qiao Y, Choi J, et al. Enhanced Association of Tau Pathology and Cognitive Impairment in Mild Cognitive Impairment Subjects with Behavior Symptoms. J Alzheimers Dis. 2022;87(2):557–568. doi:10.3233/JAD-215555

37. Dumurgier J, Sabia S, Zetterberg H, et al. A Pragmatic, Data-Driven Method to Determine Cutoffs for CSF Biomarkers of Alzheimer Disease Based on Validation Against PET Imaging. Neurology. Aug 16 2022;99(7):e669–e678. doi:10.1212/WNL.0000000000200735

38. Bates D, Mächler M, Bolker B, Walker S. Fitting linear mixed-effects models using lme4. arXiv preprint arXiv:14065823. 2014;

39. Lüdecke D. ggeffects: Tidy data frames of marginal effects from regression models. Journal of open source software. 2018;3(26):772.

40. Kaup AR, Nettiksimmons J, LeBlanc ES, Yaffe K. Memory complaints and risk of cognitive impairment after nearly 2 decades among older women. Neurology. 2015/11/24/ 2015;85(21):1852–1858. doi:10.1212/WNL.0000000000002153

41. Verlinden VJA, van der Geest JN, de Bruijn RFAG, Hofman A, Koudstaal PJ, Ikram MA. Trajectories of decline in cognition and daily functioning in preclinical dementia. Alzheimers Dement. Feb 2016;12(2):144–153. doi:10.1016/j.jalz.2015.08.001

42. Hanseeuw BJ, Scott MR, Sikkes SAM, et al. Evolution of anosognosia in alzheimer’s disease and its relationship to amyloid. Ann Neurol. 02 2020;87(2):267–280. doi:10.1002/ana.25649

43. Sperling RA, Donohue MC, Raman R, et al. Association of Factors With Elevated Amyloid Burden in Clinically Normal Older Individuals. JAMA Neurol. Jun 2020;77(6):735–745. doi:10.1001/jamaneurol.2020.0387

44. Zwan MD, Villemagne VL, Doré V, et al. Subjective Memory Complaints in APOEL4 Carriers are Associated with High Amyloid-β Burden. Journal of Alzheimer’s disease: JAD. 2016 2016;49(4):1115–1122. doi:10.3233/JAD-150446

45. Verfaillie SCJ, Timmers T, Slot RER, et al. Amyloid-β Load Is Related to Worries, but Not to Severity of Cognitive Complaints in Individuals With Subjective Cognitive Decline: The SCIENCe Project. Frontiers in Aging Neuroscience. 2019/01/25/ 2019;11doi:10.3389/fnagi.2019.00007

46. Hollands S, Lim YY, Buckley R, et al. Amyloid-β related memory decline is not associated with subjective or informant rated cognitive impairment in healthy adults. Journal of Alzheimer’s disease: JAD. 2015 2015;43(2):677–686. doi:10.3233/JAD-140678

47. Kai S. Amyloid and SCD jointly predict cognitive decline across Chinese and German cohorts. 2024.

48. Kuhn E, Klinger HM, Amariglio RE, et al. SCD-plus features and AD biomarkers in cognitively unimpaired samples: A meta-analytic approach for nine cohort studies. Alzheimers Dement. Feb 22 2025;doi:10.1002/alz.14307

49. Shao K, Hu X, Kleineidam L, et al. Amyloid and SCD jointly predict cognitive decline across Chinese and German cohorts. Alzheimers Dement. Sep 2024;20(9):5926–5939. doi:10.1002/alz.14119

50. Sanchez-Benavides G, Salvado G, Arenaza-Urquijo EM, et al. Quantitative informant- and self-reports of subjective cognitive decline predict amyloid beta PET outcomes in cognitively unimpaired individuals independently of age and APOE epsilon4. Alzheimers Dement (Amst). 2020;12(1):e12127. doi:10.1002/dad2.12127

51. Kuhn E, Perrotin A, La Joie R, et al. Association of the Informant-Reported Memory Decline With Cognitive and Brain Deterioration Through the Alzheimer Clinical Continuum. Neurology. 2023/06/13/ 2023;100(24):e2454–e2465. doi:10.1212/WNL.0000000000207338

52. Dubois B, Villain N, Frisoni GB, et al. Clinical diagnosis of Alzheimer’s disease: recommendations of the International Working Group. Lancet Neurol. 06 2021;20(6):484–496. doi:10.1016/S1474-4422(21)00066-1

