## Supplementary Online Content for "Subjective cognition trajectories, Alzheimer biomarkers, and incident mild cognitive impairment"

### eMethods. Participants and Study Design, clinical follow-up data, and AD biomarkers.

#### 1. DELCODE (DZNE-Longitudinal Cognitive Impairment and Dementia Study).

##### 1.1. Participants:

The DELCODE study is a longitudinal, observational, memory clinic-based study conducted across nine sites in Germany, seven of which are linked to local university memory centers. Initiated in 2014 under the leadership of Prof. Dr. med. Frank Jessen, inclusion and exclusion criteria are described in Jessen et al., 2018.1 The current study focuses on controls, first-degree relatives of Alzheimer's disease (AD) patients, and patients with subjective cognitive decline (SCD). Recruitment occurred through local newspaper advertisements and memory center referrals. All participants scored better than 1·5 standard deviations below the age-, sex-, and education-adjusted norms on the CERAD neuropsychological battery (German norms available at www.memoryclinic.ch). SCD patients were additionally defined by subjectively reported cognitive decline, with concerns raised to memory center physicians, in line with criteria from previous studies.2,3 Participants in the current study were aged 60-84, had at least 8 years of education, and no significant psychiatric, neurological, or substance abuse history. They also did not take psychoactive or anti-dementia medications. In total, 151 controls, 60 AD-relatives, and 279 SCD patients were included. They were enrolled between May 13, 2014, and August 31, 2018.

##### 1.2. Clinical conversion to incident-MCI:

Consensus diagnoses of incident MCI were determined through a two-step review process, adapted from the Wisconsin Registry for Alzheimer's Prevention study and described in Stark et al, 2023.[4] In the first step, an algorithmic screening identified participants who were cognitively normal at baseline but showed potential cognitive decline at follow-up. The criteria for this included: (i) normative deficits (e.g., at least two cognitive tests ≥1·0 SD below the demographically adjusted mean, or one CERAD-NAB subtest ≥1·5 SD below the mean), (ii) individual cut-offs (e.g., CERAD-NAB word list delayed recall or FCSRT free recall ≥1·5 SD below the mean, FCSRT total recall ≤46 points, or MMSE ≤26 points), and (iii) reports from physicians or study partners (e.g., an increase in CDR global score, or FAQ score ≥4). In the second step, flagged cases were reviewed by a team of five neuropsychologists who assessed MCI conversion using established diagnostic criteria: (i) cognitive impairment (≥1·5 SD below the mean in at least two tests measuring the same ability), (ii) intra-individual cognitive decline (based on longitudinal test scores or participant/study partner reports), (iii) preserved functional abilities, and (iv) absence of dementia.[5,6] The review committee was blinded to baseline group assignments, biomarkers, imaging data, and genetic information.

##### 1.3. AD biomarkers:

In the current study, amyloid and tau positivity were first assessed using cerebrospinal fluid (CSF) data, and plasma data when CSF was unavailable.
**CSF Biomarkers**: A subset of participants underwent lumbar puncture for CSF sampling (N=274, sampling rate: 55·9%). Amyloid beta (Aβ)42, Aβ40, and phospho-tau181 (p-tau181) levels were measured using standardized commercial kits: V-PLEX Aβ Peptide Panel 1 (6E10) Kit (K15200E, Mesoscale Diagnostics LLC, USA) and Innotest Phospho-Tau(181P) (81581; Fujirebio Germany GmbH). Assays were performed centrally, with independent reference samples used for quality control. Cut-offs were derived using Gaussian mixture modeling (R package flexmix, version 2.3-15)[7] and set at Aβ42/40 ≤ 0·08 and p-tau181 ≥ 73·65 pg/ml.[8]
**Plasma Aβ42/40:** Plasma samples (500 µL EDTA) were processed according to DELCODE standard operating procedures and stored at −80°C. AβX-40 and AβX-42 levels, along with the AβX-42/X-40 ratio, were measured using a semi-automated immunoprecipitation-immunoassay (IP-IA). The plasma Aβ immunoprecipitation was performed using a CyBio FeliX liquid-handling instrument (Roboscreen, Leipzig, Germany), followed by detection with the Mesoscale Discovery Aβ V-PLEX immunoassay (6E10). The cut-off for plasma Aβ42/40 was set at ≤ 0·106, as detailed in Vogelgsang et al., 2024.[9]
**Plasma pTau181**: Plasma levels of p-tau181 were measured using Simoa assays on Quanterix HD-1 and HD-X instruments (Quanterix, Billerica, MA) with pTau-181 Advantage version 2 kits, following the manufacturer's instructions. Assays were performed by the same operator, with two internal control samples assessed at the start and end of each run to ensure repeatability and monitor inter-assay variability.[10] The plasma p-tau181 cut-off for this study was determined via receiver operating characteristic (ROC) analysis, using CSF p-tau181 classification and the Youden Index to establish the threshold. The cut-off was set at ≥1·707 pg/mL, with an area under the curve (AUC) of 0·78 (95% CI: 0·73-0·83), based on 466 DELCODE participants, including MCI and demented patients (unpublished).

#### 2. ADNI (Alzheimer's Disease Neuroimaging Initiative).

In the current study, we utilized data from the ADNIMERGE dataset, extracted on September 21, 2023.

##### 2.1. Participants:

ADNI is an ongoing, longitudinal, multicenter study conducted in 63 sites across the USA and Canada. The ADNI was launched in 2003 as a public-private partnership, led by Principal Investigator Michael W. Weiner, MD. The primary goal of ADNI has been to test whether serial magnetic resonance imaging (MRI), positron emission tomography (PET), other biological markers, and clinical and neuropsychological assessment can be combined to measure the progression of mild cognitive impairment (MCI) and early Alzheimer's disease (AD). Further details can be found in Weiner et al,[11] and inclusion and exclusion criteria have been described at http://adni.loni.usc.edu/. Briefly, participants included in the current study were all aged between 56-90 years, had at least 8 years of education, and had no clinically significant psychiatric (including alcohol or drug abuse) or neurologic disease. All cognitively unimpaired (CU) participants had MMSE[12] scores of 24 or greater (total possible range: 0-30) and Clinical Dementia Rating (CDR) of 0.0 (total possible range 0-3, with higher scores indicating worse functioning)[13], are not depressed, did not have mild cognitive impairment or dementia, have preserved activities of daily living (FAQ≤9), and had scores on delayed recall of one paragraph from Wechsler Memory Scale Logical Memory II within the norm according to their level of education (≥9 for 16 or more years of education, ≥5 for 8-15 years of education, ≥3 for 0-7 years of education). Part of those participants (N=145 [51·8%]) had a significant subjective cognitive decline, defined by the presence of a significant subjective memory concern reported by subject, informant, or clinician, CCI score ≥16 (based on first 12 questions).[14]

**Clinical Conversion to Incident MCI**: To determine the presence of a clinical conversion during the follow-up period, we used an algorithm based on the "DX" (diagnosis) column from the ADNIMERGE dataset. Individuals diagnosed with mild cognitive impairment (MCI) at any point during follow-up were considered converters to incident MCI from the time of diagnosis. However, individuals with reversible MCI, defined as those who had an inconsistent MCI diagnosis, followed by a return to normal cognition in subsequent assessments (with at least three total assessments), were considered stable and not counted as converters.

##### 2.2. AD biomarkers:

**Ab-PET biomarkers:** The ADNI FreeSurfer 5.3 pipeline yields a global standard uptake value ratio (SUVR) measure representing a non-weighted average of radiotracer retention in four FreeSurfer-defined regions (frontal, anterior/posterior cingulate, lateral parietal, and lateral temporal cortices) normalized to whole cerebellum. To directly convert [18F]-Florbetapir- and [18F]-Florbetaben-PET data to centiloid values, equations describe in ADNI guidelines were used (see details here). Centiloids were calculated as follows:

- [18F]-Florbetapir-PET: CL=(196·9×SUVR)-196·03. The amyloid positivity threshold was set at 22·529, corresponding to 1·11 SUVR (see also UCBERKELEY_AV45_Methods_11.15.2021.pdf).[15,16]
- [18F]-Florbetaben-PET: CL=(159·08×SUVR)-151·65. The amyloid positivity threshold was set at 20·1564, corresponding to 1·08 SUVR (see also UCBerkeley_FBB_Methods_11.15.2021.pdf).[17]

**Tau-PET biomarker:** The processing of [18F]-Flortaucipir PET images is summarized as follows (please see here for details). Each PET scan is co-registered to the closest available bias-corrected T1-weighted MRI in native space, using FreeSurfer (v7.1.1) for anatomical accuracy. This registration allows the calculation of the mean flortaucipir uptake within predefined regions of interest (ROIs). For the current study, tau positivity is defined based on the standardized uptake value ratio (SUVR) in the entorhinal cortex (corresponding to Braak stage 1). A positivity threshold of SUVR≥1·2 was used, as established in a previously published paper.[18]

**CSF ptau181:** Data were extracted from the "PTAU" variable in the ADNIMERGE dataset, that use Roche Elecsys immunoassays. A positivity threshold of ≥24·3 pg/mL was used, as established in a previously published paper,[19] which reported an area under the curve (AUC) of 0·79 ± 0·03.

**eTable 1.** Baseline participants demographics by cohort subsample

| | DELCODE (N=490) | | | | | | ADNI (N=280) | | | | | |
|---|---|---|---|---|---|---|---|---|---|---|---|---|
| | Ab-Stable | Ab+Stable | Ab-iMCI | Ab+iMCI | Overall | P | Ab-Stable | Ab+Stable | Ab-iMCI | Ab+iMCI | Overall | P |
| No. (%) | 290 (59·2%) | 104 (21·2%) | 53 (10·8%) | 43 (8·8%) | 490 (100%) | | 172 (61·4%) | 73 (26·1%) | 16 (5·7%) | 19 (6·8%) | 280 (100%) | |
| FU time, median (IQR), y[a] | 5·1 (4·0-6·9) | 5·0 (4·0-7·0) | 4·2 (3·4-6·0) | 5·0 (4·0-6·1) | 5·1 (4·0-6·6) | *·05* | 4·4 (3·2-7·7) | 4·0 (2·6-5·3) | 6·9 (4·2-8·0) | 4·0 (2·5-5·4) | 4·2 (3·0-7·5) | **·02** |
| Age, median (IQR), y[b] | 68·0 (64·3-72·4) | 72·1 (67·5-76·4) | 71·0 (67·3-76·4) | 74·4 (69·2-77·5) | 69·8 (65·3-74·5) | **<·001** | 69·4 (66·1-74·0) | 70·4 (67·4-76·4) | 70·8 (66·8-76·1) | 75·1 (71·1-81·0) | 70·0 (66·7-75·3) | **<·001** |
| Female, No. (%)[c] | 170 (58·6%) | 30 (28·8%) | 28 (52·8%) | 15 (34·9%) | 243 (49·6%) | **<·001** | 97 (56·4%) | 49 (67·1%) | 7 (43·8%) | 9 (47·4%) | 162 (57·9%) | ·18 |
| Education, median (IQR), y | 15 (13-17) | 15 (13-18) | 13 (12-17) | 13 (12-17) | 14 (13-17) | ·34 | 17 (16-18) | 16 (14-19) | 16 (13·8-18·3) | 16 (16-20) | 16·5 (16-18) | ·46 |
| APOEe4 carrier, No. (%)[d] | 45 (15·6%) | 54 (52·4%) | 13 (24·5%) | 23 (53·5%) | 135 (27·7%) | **<·001** | 40 (24·1%) | 40 (56·3%) | 3 (18·8%) | 10 (55·6%) | 93 (34·3%) | **<·001** |
| MMSE score, median (IQR)[e] | 30 (29-30) | 30 (29-30) | 29 (28-30) | 29 (28·5-30) | 30 (29-30) | **<·001** | 29 (29-30) | 29 (29-30) | 29 (27-30) | 29 (28-30) | 29 (29-30) | ·11 |
| Memory clinic, No. (%)[f] | 148 (51·0%) | 60 (57·7%) | 36 (67·9%) | 35 (81·4%) | 279 (56·9%) | **<·001** | | | | | | |
| No. (%) with tau status | 270 (59·5%) | 93 (20·5%) | 53 (11·7%) | 38 (8·4%) | 454 (100%) | | 157 (61·3%) | 71 (27·7%) | 12 (4·7%) | 16 (6·3%) | 256 (100%) | |
| Tau status T+, No. (%)[fg] | 18 (6·7%) | 18 (19·4%) | 7 (13·2%) | 15 (39·5%) | 58 (12·8%) | **<·001** | 27 (17·2%) | 34 (47·9%) | 5 (41·7%) | 10 (62·5%) | 76 (29·7%) | **<·001** |

At Bonferroni-adjusted P values (for 6 intergroup comparisons), in DELCODE: [b] Ab-Stable younger than the 3 others groups (all $P_{adj}$<0·01). [c,d] Both Ab- groups with higher percentage of female than Ab+ (all $P_{adj}$<0·04) and lower percentage of APOE4 carriers (all $P_{adj}$<0·04), except for Ab-iMCI and Ab+iMCI which did not significantly differ in sex ratio ($P_{adj}$=0·72). Note that only 488 DELCODE participants had available APOE data (1 missing both Stable groups). [e] Both Stable groups with higher MMSE scores than both iMCI groups (all $P_{adj}$<0·04), with no significant differences based on Ab status (both $P_{adj}$=1). [f] Ab+iMCI had a higher percentage of SCD patients compared to Ab-Stable ($P_{adj}$=0·002). [g] Ab+iMCI had a higher percentage of tau-positive individuals compared to the Ab- groups (all $P_{adj}$<0·04), as Ab+Stable compared to Ab-Stable group ($P_{adj}$<0·001). In ADNI: [a] Ab-iMCI with longer follow-up period than Ab+Stable ($P_{adj}$=0·02). [b] Ab+iMCI older than both Stable groups (both $P_{adj}$<0·03). [d] Both Ab- groups with lower percentage of APOE4 carriers compared to both Ab+ groups (between both Stable groups, $P_{adj}$<0·001; only trend level for others, both $P_{adj}$<0·08), except for Ab-iMCI and Ab+iMCI which did not significantly differ ($P_{adj}$=0·36). Note that only 271 ADNI participants had available APOE data (6 missing in Ab-Stable, 2 missing in Ab+Stable and 1 mising in Ab+iMCI). [g] Ab-Stable had a lower percentage of tau-positive individuals compared both Ab+ groups (all $P_{adj}$<0·001). Abbreviations: Ab-, participants amyloid-negative at baseline; Ab+, participants amyloid-positive at baseline; ADNI, Alzheimer's Disease Neuroimaging Initiative; FU, Follow-up, iMCI, participants that progressed to incident mild cognitive impairment during the follow-up period; DELCODE, German Center for Neurodegenerative Diseases (DZNE) Longitudinal Cognitive Impairment and Dementia Study; MMSE, mini mental state examination; Stable, participants cognitively stable during the follow-up period; T+, participants tau-positive at baseline.

**eTable 2.** Participants demographics at the first follow-up (6-18 months after baseline) for the subset with DC1 scores available that did not yet convert to MCI

| | Combined Sample (N=353) | | | | | |
|---|---|---|---|---|---|---|
| | Ab-Stable | Ab+Stable | Ab-iMCI | Ab+iMCI | Overall | P value |
| No. (%) | 204 (57·8%) | 97 (27·5%) | 26 (7·4%) | 26 (7·4%) | 353 (100%) | |
| DELCODE, No. (%) | 123 (60·3%) | 54 (55·7%) | 20 (76·9%) | 18 (69·2%) | 215 (60·9%) | |
| Memory clinic[a], No. (%) | 81 (39·7%) | 33 (34·0%) | 15 (57·7%) | 14 (53·8%) | 143 (40·5%) | **<0·001** |
| Age[b], median (IQR), y | 69·4 (65·7-73·9) | 72·2 (68·2-76·7) | 72·5 (70·8-78·6) | 76·3 (73·1-80·0) | 71·0 (67·1-75·9) | **<0·001** |
| Female[c], No. (%) | 10 (53·9%) | 38 (39·2%) | 9 (34·6%) | 11 (42·3%) | 168 (47·6%) | **0·04** |
| Education[d], median (IQR), y | 16 (13-18) | 16 (14-18) | 15 (13-17) | 14 (12-16) | 16 (13-18) | **0·01** |
| APOEe4 carrier[e], No. (%) | 37 (18·4%) | 53 (56·4%) | 6 (23·1%) | 14 (53·8%) | 110 (31·7%) | **<0·001** |
| MMSE score[f], median (IQR) | 30 (29-30) | 30 (28-30) | 29 (28-30) | 29 (27·3-30) | 30 (29-30) | **0·003** |
| No. (%) with tau status | 188 (57·3%) | 93 (28·4%) | 25 (7·6%) | 22 (6·7%) | 328 (100%) | |
| Tau status T+[g], No. (%) | 19 (10·1%) | 36 (38·7%) | 7 (28·0%) | 9 (40·9%) | 71 (21·6%) | **<0·001** |

At Bonferroni-adjusted P values (for 6 intergroup comparisons): [a] iMCI groups have more participants recruited from memory clinics than Stable groups (all $P_{adj}$<0·03). [b,g] Ab-Stable younger than the Ab+ groups (all $P_{adj}$<0·003) and with lower percentage of tau-positive participants (all $P_{adj}$≤0·001). [c] No significant differences between groups. [d] Ab+Stable more educated than the Ab-iMCI group (all $P_{adj}$<0·01). [e] Only 347 participants (3 missing in Ab-Stable, and 3 missing in Ab+Stable) had available APOE data. Ab-Stable with a lower percentage of APOEe4 carriers than the Ab+ groups (all $P_{adj}$<0·001), and Ab-iMCI than Ab+Stable ($P_{adj}$=0·03). [f] Only 345 participants (2 missing in Ab-Stable, and 6 missing in Ab+Stable) had available MMSE data. Both Stable groups with higher MMSE scores than Ab+iMCI groups (all $P_{adj}$<0·02). Abbreviations: Ab-, participants amyloid-negative at baseline; Ab+, participants amyloid-positive at baseline; FU, Follow-up, iMCI, participants that progressed to incident mild cognitive impairment during the follow-up period; MMSE, mini mental state examination; Stable, participants cognitively stable during the follow-up period; T+, participants tau-positive at baseline.

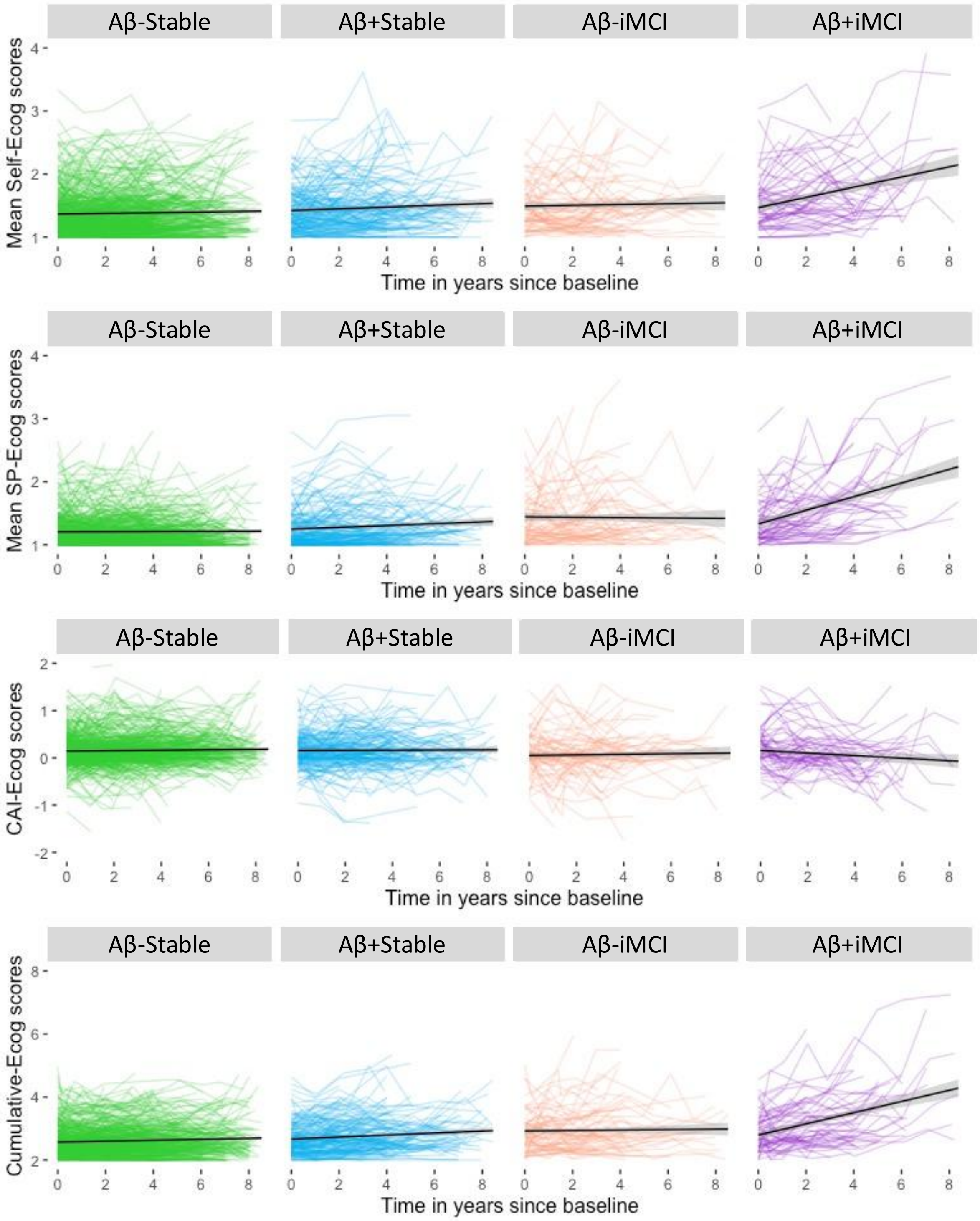

**eFigure 1.** Spaghetti plot of individual trajectories: SCD reports over time in years since baseline according to amyloid status and future clinical progression.

Abbreviations: Ab-, participants amyloid-negative at baseline; Ab+, participants amyloid-positive at baseline; CAI, Cognitive awareness index; iMCI, participants that progressed to incident mild cognitive impairment during the follow-up period; SP, study partner; Stable, participants cognitively stable during the follow-up period.

**eTable 3.** Results from Models 1-3, including subgroup slopes and pairwise comparisons with Bonferroni-adjusted p-values using the ggeffects::hypothesis_test() function.

**eTable 4A.** Combined sample.

| | **SP-Ecog** | | | **Self-Ecog** | | | **Cum-Ecog** | | | **CAI-Ecog** | | |
|---|---|---|---|---|---|---|---|---|---|---|---|---|
| **Model 1 - Fixed Effect[a]** | Est. | SE | P | Est. | SE | P | Est. | SE | P | Est. | SE | P |
| (Intercept) | 1·18 | 0·02 | **<0·001** | 1·21 | 0·03 | **<0·001** | 2·38 | 0·04 | **<0·001** | 0·02 | 0·03 | 0·43 |
| Time | 0·01 | 0·004 | **0·006** | 0·02 | 0·005 | **<0·001** | 0·03 | 0·007 | **<0·001** | 0·006 | 0·005 | 0·22 |
| iMCI [iMCI] | 0·13 | 0·03 | **<0·001** | 0·08 | 0·03 | **0·012** | 0·21 | 0·04 | **<0·001** | -0·05 | 0·03 | 0·18 |
| Time:iMCI [iMCI] | 0·04 | 0·006 | **<0·001** | 0·02 | 0·006 | **<0·001** | 0·06 | 0·009 | **<0·001** | -0·01 | 0·006 | *0·02* |
| **Slopes** | Est. | 95% CI | $P_{adj}$ | Est. | 95% CI | $P_{adj}$ | Est. | 95% CI | $P_{adj}$ | Est. | 95% CI | $P_{adj}$ |
| Stable | 0·01 | 0·003, 0·02 | **0·02** | 0·02 | 0·006, 0·02 | **0·002** | 0·03 | 0·01, 0·04 | **<0·001** | 0·006 | -0·003, 0·01 | 0·41 |
| iMCI | 0·05 | 0·04, 0·07 | **<0·001** | 0·04 | 0·02, 0·05 | **<0·001** | 0·09 | 0·07, 0·11 | **<0·001** | -0·008 | -0·02, 0·006 | 0·48 |
| **Model 2 - Fixed Effect[b]** | Est. | SE | P | Est. | SE | P | Est. | SE | P | Est. | SE | P |
| (Intercept) | 1·19 | 0·03 | **<0·001** | 1·23 | 0·03 | **<0·001** | 2·41 | 0·05 | **<0·001** | 0·03 | 0·04 | 0·45 |
| Time | 0·007 | 0·006 | 0·25 | 0·01 | 0·006 | *0·04* | 0·02 | 0·009 | *0·03* | 0·007 | 0·006 | 0·26 |
| Amyloid status [Ab+] | 0·009 | 0·02 | 0·67 | 0·04 | 0·03 | 0·17 | 0·04 | 0·04 | 0·26 | 0·02 | 0·03 | 0·49 |
| Time:Amyloid status [Ab+] | 0·03 | 0·005 | **<0·001** | 0·02 | 0·005 | **<0·001** | 0·04 | 0·007 | **<0·001** | -0·006 | 0·005 | 0·21 |
| **Slopes** | Est. | 95% CI | $P_{adj}$ | Est. | 95% CI | $P_{adj}$ | Est. | 95% CI | $P_{adj}$ | Est. | 95% CI | $P_{adj}$ |
| Ab- | 0·006 | -0·005, 0·02 | 0·62 | 0·01 | -0·0000, 0·02 | 0·10 | 0·02 | 0·001, 0·04 | *0·07* | 0·007 | -0·005, 0·02 | 0·49 |
| Ab+ | 0·03 | 0·02, 0·05 | **<0·001** | 0·03 | 0·02, 0·05 | **<0·001** | 0·06 | 0·04, 0·08 | **<0·001** | 0·0007 | -0·01, 0·01 | 1·00 |
| **Model 3 - Fixed Effect[b]** | Est. | SE | P | Est. | SE | P | Est. | SE | P | Est. | SE | P |
| (Intercept) | 1·17 | 0·03 | **<0·001** | 1·22 | 0·03 | **<0·001** | 2·38 | 0·05 | **<0·001** | 0·04 | 0·04 | 0·31 |
| Time | 0·005 | 0·006 | 0·33 | 0·01 | 0·006 | *0·07* | 0·02 | 0·009 | *0·05* | 0·007 | 0·006 | 0·29 |
| iMCI [iMCI] | 0·17 | 0·03 | **<0·001** | 0·07 | 0·04 | *0·09* | 0·24 | 0·06 | **<0·001** | -0·08 | 0·04 | *0·06* |
| Amyloid status [Ab+] | 0·01 | 0·02 | 0·53 | 0·03 | 0·03 | 0·34 | 0·03 | 0·04 | 0·38 | 0·009 | 0·03 | 0·74 |
| Time:iMCI [iMCI] | 0·01 | 0·007 | *0·07* | 0·01 | 0·008 | 0·13 | 0·02 | 0·01 | *0·05* | 0·0001 | 0·008 | 0·99 |
| Time:Amyloid status [Ab+] | 0·01 | 0·005 | *0·02* | 0·02 | 0·006 | **0·006** | 0·03 | 0·008 | **<0·001** | 0·001 | 0·005 | 0·83 |
| iMCI:Amyloid status [Ab+iMCI] | -0·09 | 0·05 | *0·08* | 0·006 | 0·06 | 0·93 | -0·08 | 0·09 | 0·38 | 0·08 | 0·07 | 0·25 |
| Time:iMCI:Amyloid status [Ab+iMCI] | 0·06 | 0·01 | **<0·001** | 0·02 | 0·01 | 0·19 | 0·07 | 0·02 | **<0·001** | -0·03 | 0·01 | **0·009** |
| **Slopes** | Est. | 95% CI | $P_{adj}$ | Est. | 95% CI | $P_{adj}$ | Est. | 95% CI | $P_{adj}$ | Est.Est. | 95% CI | $P_{adj}$ |
| Ab-Stable | 0·005 | -0·006, 0·02 | 1·00 | 0·01 | -0·001, 0·02 | 0·32 | 0·02 | -0·0007,0·03 | 0·24 | 0·007 | -0·005, 0·02 | 1·00 |
| Ab+Stable | 0·02 | 0·005, 0·03 | **0·03** | 0·03 | 0·01, 0·04 | **0·001** | 0·04 | 0·02, 0·06 | **<0·001** | 0·008 | -0·006, 0·02 | 1·00 |
| Ab-iMCI | 0·02 | 0·002, 0·04 | 0·13 | 0·02 | 0·004, 0·04 | *0·06* | 0·04 | 0·01, 0·06 | **0·01** | 0·007 | -0·01, 0·03 | 1·00 |
| Ab+iMCI | 0·09 | 0·07, 0·11 | **<0·001** | 0·05 | 0·03, 0·07 | **<0·001** | 0·13 | 0·11, 0·16 | **<0·001** | -0·02 | -0·04, -0·004 | *0·07* |
| **Pairwise comparisons** | Est. | 95% CI | $P_{adj}$ | Slope | 95% CI | $P_{adj}$ | Slope | 95% CI | $P_{adj}$ | Slope | 95% CI | $P_{adj}$ |
| Ab-Stable / Ab+Stable | -0·01 | -0·02, -0·002 | *0·09* | -0·02 | -0·03, -0·004 | **0·04** | -0·03 | -0·04, -0·01 | **0·005** | -0·001 | -0·01, 0·01 | 1·00 |
| Ab-Stable / Ab-iMCI | -0·01 | -0·03, 0·001 | 0·40 | -0·01 | -0·03, 0·004 | 0·80 | -0·02 | -0·04,-0·0003 | 0·28 | -0·0001 | -0·02, 0·02 | 1·00 |
| Ab-Stable / Ab+iMCI | -0·08 | -0·10, -0·07 | **<0·001** | -0·04 | -0·06, -0·03 | **<0·001** | -0·12 | -0·14, -0·09 | **<0·001** | 0·03 | 0·01, 0·05 | **0·003** |
| Ab+Stable / Ab-iMCI | -0·0007 | -0·02, 0·02 | 1·00 | 0·003 | -0·01, 0·02 | 1·00 | 0·003 | -0·02, 0·03 | 1·00 | 0·001 | -0·02, 0·02 | 1·00 |
| Ab+Stable / Ab+iMCI | -0·07 | -0·09, -0·05 | **<0·001** | -0·03 | -0·05, -0·009 | **0·02** | -0·09 | -0·12, -0·07 | **<0·001** | 0·03 | 0·01, 0·05 | **0·004** |
| Ab-iMCI / Ab+iMCI | -0·07 | -0·09, -0·05 | **<0·001** | -0·03 | -0·05, -0·01 | **0·02** | -0·10 | -0·13, -0·07 | **<0·001** | 0·03 | 0·009, 0·05 | **0·03** |

**eTable 5B.** DELCODE cohort.

| | **SP-Ecog** | | | **Self-Ecog** | | | **Cum-Ecog** | | | **CAI-Ecog** | | |
|---|---|---|---|---|---|---|---|---|---|---|---|---|
| **Model 1 - Fixed Effect[a]** | Est. | SE | P | Est. | SE | P | Est. | SE | P | Est. | SE | P |
| (Intercept) | 1·20 | 0·02 | **<0·001** | 1·22 | 0·03 | **<0·001** | 2·41 | 0·04 | **<0·001** | 0·01 | 0·03 | 0·71 |
| Time | 0·01 | 0·004 | **0·001** | 0·02 | 0·005 | **0·001** | 0·03 | 0·007 | **<0·001** | 0·005 | 0·005 | 0·37 |
| iMCI [iMCI] | 0·11 | 0·03 | **<0·001** | 0·05 | 0·04 | 0·16 | 0·17 | 0·05 | **<0·001** | -0·05 | 0·04 | 0·26 |
| Time:iMCI [iMCI] | 0·04 | 0·006 | **<0·001** | 0·03 | 0·007 | **<0·001** | 0·07 | 0·01 | **<0·001** | -0·01 | 0·008 | 0·12 |
| **Slopes** | Est. | 95% CI | $P_{adj}$ | Est. | 95% CI | $P_{adj}$ | Est. | 95% CI | $P_{adj}$ | Est. | 95% CI | $P_{adj}$ |
| Stable | 0·01 | 0·005, 0·02 | **0·003** | 0·02 | 0·006, 0·02 | **0·002** | 0·03 | 0·02, 0·05 | **<0·001** | 0·005 | -0·006, 0·01 | 0·75 |
| iMCI | 0·05 | 0·04, 0·07 | **<0·001** | 0·04 | 0·03, 0·06 | **<0·001** | 0·10 | 0·08, 0·12 | **<0·001** | -0·008 | -0·02, 0·009 | 0·73 |
| **Model 2 - Fixed Effect[b]** | Est. | SE | P | Est. | SE | P | Est. | SE | P | Est. | SE | P |
| (Intercept) | 1·22 | 0·03 | **<0·001** | 1·24 | 0·03 | **<0·001** | 2·45 | 0·04 | **<0·001** | 0·02 | 0·04 | 0·65 |
| Time | 0·01 | 0·005 | *0·05* | 0·01 | 0·006 | *0·02* | 0·03 | 0·009 | **0·003** | 0·007 | 0·006 | 0·23 |
| Amyloid status [Ab+] | -0·01 | 0·03 | 0·62 | 0·009 | 0·03 | 0·78 | -0·008 | 0·04 | 0·85 | 0·02 | 0·04 | 0·66 |
| Time:Amyloid status [Ab+] | 0·03 | 0·005 | **<0·001** | 0·02 | 0·006 | **0·004** | 0·05 | 0·01 | **<0·001** | -0·02 | 0·007 | **0·009** |
| **Slopes** | Est. | 95% CI | $P_{adj}$ | Est. | 95% CI | $P_{adj}$ | Est. | 95% CI | $P_{adj}$ | Est. | 95% CI | $P_{adj}$ |
| Ab- | 0·01 | 0·0003, 0·02 | 0·11 | 0·01 | 0·002, 0·02 | **0·046** | 0·03 | 0·008, 0·04 | **0·009** | 0·007 | -0·005, 0·02 | 0·47 |
| Ab+ | 0·04 | 0·03, 0·05 | **<0·001** | 0·03 | 0·02, 0·04 | **<0·001** | 0·08 | 0·06, 0·10 | **<0·001** | -0·01 | -0·02, 0·004 | 0·33 |
| **Model 3 - Fixed Effect[b]** | Est. | SE | P | Est. | SE | P | Est. | SE | P | Est. | SE | P |
| (Intercept) | 1·20 | 0·03 | **<0·001** | 1·24 | 0·03 | **<0·001** | 2·42 | 0·04 | **<0·001** | 0·02 | 0·04 | 0·54 |
| Time | 0·01 | 0·005 | *0·04* | 0·01 | 0·006 | *0·05* | 0·03 | 0·008 | **0·004** | 0·006 | 0·006 | 0·31 |
| iMCI [iMCI] | 0·12 | 0·04 | **0·002** | 0·04 | 0·05 | 0·37 | 0·18 | 0·06 | **0·005** | -0·06 | 0·06 | 0·32 |
| Amyloid status [Ab+] | -0·03 | 0·03 | 0·45 | 0·0000 | 0·04 | 1·00 | -0·02 | 0·05 | 0·66 | 0·02 | 0·04 | 0·58 |
| Time:iMCI [iMCI] | 0·007 | 0·008 | 0·38 | 0·02 | 0·009 | *0·06* | 0·02 | 0·01 | 0·10 | 0·005 | 0·01 | 0·60 |
| Time:Amyloid status [Ab+] | 0·01 | 0·006 | **0·011** | 0·01 | 0·007 | 0·15 | 0·02 | 0·01 | *0·03* | -0·009 | 0·007 | 0·24 |
| iMCI:Amyloid status [Ab+iMCI] | -0·02 | 0·06 | 0·77 | 0·02 | 0·07 | 0·82 | -0·03 | 0·10 | 0·80 | -0·0000 | 0·09 | 1·00 |
| Time:iMCI:Amyloid status [Ab+iMCI] | 0·07 | 0·01 | **<0·001** | 0·02 | 0·01 | 0·11 | 0·10 | 0·02 | **<0·001** | -0·04 | 0·02 | *0·02* |
| **Slopes** | Est. | 95% CI | $P_{adj}$ | Est. | 95% CI | $P_{adj}$ | Est. | 95% CI | $P_{adj}$ | Est. | 95% CI | $P_{adj}$ |
| Ab-Stable | 0·01 | 0·0004, 0·02 | 0·16 | 0·01 | -0·0000, 0·02 | 0·20 | 0·02 | 0·007, 0·04 | **0·02** | 0·006 | -0·006, 0·02 | 1·00 |
| Ab+Stable | 0·02 | 0·01, 0·04 | **<0·001** | 0·02 | 0·007, 0·03 | **0·01** | 0·05 | 0·03, 0·07 | **<0·001** | -0·002 | -0·02, 0·01 | 1·00 |
| Ab-iMCI | 0·02 | 0·0004, 0·03 | 0·18 | 0·03 | 0·009, 0·05 | **0·02** | 0·05 | 0·02, 0·07 | **0·006** | 0·01 | -0·01, 0·03 | 1·00 |
| Ab+iMCI | 0·10 | 0·08, 0·12 | **<0·001** | 0·06 | 0·04, 0·08 | **<0·001** | 0·17 | 0·14, 0·20 | **<0·001** | -0·03 | -0·06, -0·01 | **0·02** |
| **Pairwise comparisons** | Est. | 95% CI | $P_{adj}$ | Slope | 95% CI | $P_{adj}$ | Slope | 95% CI | $P_{adj}$ | Slope | 95% CI | $P_{adj}$ |
| Ab-Stable / Ab+Stable | -0·01 | -0·03, -0·003 | *0·07* | -0·01 | -0·02, 0·003 | 0·88 | -0·02 | -0·04, -0·003 | 0·16 | 0·009 | -0·006, 0·02 | 1·00 |
| Ab-Stable / Ab-iMCI | -0·007 | -0·02, 0·008 | 1·00 | -0·02 | -0·03, 0·001 | 0·39 | -0·02 | -0·05, 0·004 | 0·61 | -0·005 | -0·03, 0·01 | 1·00 |
| Ab-Stable / Ab+iMCI | -0·09 | -0·11, -0·07 | **<0·001** | -0·05 | -0·07, -0·03 | **<0·001** | -0·14 | -0·17, -0·11 | **<0·001** | 0·04 | 0·02, 0·06 | **0·003** |
| Ab+Stable / Ab-iMCI | 0·008 | -0·009, 0·02 | 1·00 | -0·007 | -0·03, 0·01 | 1·00 | -0·0000 | -0·03, 0·03 | 1·00 | -0·01 | -0·04, 0·008 | 1·00 |
| Ab+Stable / Ab+iMCI | -0·08 | -0·09, -0·06 | **<0·001** | -0·04 | -0·06, -0·02 | **0·001** | -0·12 | -0·15, -0·09 | **<0·001** | 0·03 | 0·007, 0·05 | *0·07* |
| Ab-iMCI / Ab+iMCI | -0·08 | -0·10, -0·06 | **<0·001** | -0·03 | -0·06, -0·008 | *0·06* | -0·12 | -0·16, -0·08 | **<0·001** | 0·04 | 0·02, 0·07 | **0·009** |

**eTable 3C.** ADNI cohort

| | **SP-Ecog** | | | **Self-Ecog** | | | **Cum-Ecog** | | | **CAI-Ecog** | | |
|---|---|---|---|---|---|---|---|---|---|---|---|---|
| **Model 1 - Fixed Effect[a]** | Est. | SE | P | Est. | SE | P | Est. | SE | P | Est. | SE | P |
| (Intercept) | 1·20 | 0·03 | **<0·001** | 1·39 | 0·04 | **<0·001** | 2·65 | 0·05 | **<0·001** | 0·18 | 0·03 | **<0·001** |
| Time | 0·01 | 0·009 | 0·11 | 0·02 | 0·007 | **0·003** | 0·04 | 0·008 | **<0·001** | 0·0004 | 0·005 | 0·94 |
| iMCI [iMCI] | 0·17 | 0·05 | **<0·001** | 0·14 | 0·06 | *0·03* | 0·31 | 0·08 | **<0·001** | -0·05 | 0·06 | 0·40 |
| Time:iMCI [iMCI] | 0·06 | 0·02 | **<0·001** | 0·007 | 0·01 | 0·57 | 0·04 | 0·01 | *0·02* | -0·02 | 0·01 | *0·08* |
| **Slopes** | Est. | 95% CI | $P_{adj}$ | Est. | 95% CI | $P_{adj}$ | Est. | 95% CI | $P_{adj}$ | Est. | 95% CI | $P_{adj}$ |
| Stable | 0·02 | -0·001, 0·03 | 0·15 | 0·02 | 0·007, 0·03 | **0·005** | 0·04 | 0·02, 0·05 | **<0·001** | -0·0007 | -0·01, 0·01 | 1·00 |
| iMCI | 0·07 | 0·04, 0·10 | **<0·001** | 0·03 | 0·004, 0·05 | **0·05** | 0·07 | 0·04, 0·10 | **<0·001** | -0·02 | -0·04, 0·0008 | 0·12 |
| **Model 2 - Fixed Effect[b]** | Est. | SE | P | Est. | SE | P | Est. | SE | P | Est. | SE | P |
| (Intercept) | 1·22 | 0·03 | **<0·001** | 1·40 | 0·04 | **<0·001** | 2·69 | 0·05 | **<0·001** | 0·17 | 0·03 | **<0·001** |
| Time | 0·02 | 0·009 | *0·08* | 0·01 | 0·007 | *0·03* | 0·03 | 0·008 | **<0·001** | -0·005 | 0·006 | 0·38 |
| Amyloid status [Ab+] | 0·03 | 0·03 | 0·34 | 0·07 | 0·05 | 0·12 | 0·09 | 0·06 | 0·13 | 0·04 | 0·04 | 0·37 |
| Time:Amyloid status [Ab+] | 0·02 | 0·01 | 0·11 | 0·03 | 0·009 | **0·004** | 0·03 | 0·01 | **0·001** | 0·01 | 0·008 | 0·15 |
| **Slopes** | Est. | 95% CI | $P_{adj}$ | Est. | 95% CI | $P_{adj}$ | Est. | 95% CI | $P_{adj}$ | Est. | 95% CI | $P_{adj}$ |
| Ab- | 0·02 | -0·002, 0·03 | 0·16 | 0·01 | 0·001, 0·03 | *0·06* | 0·03 | 0·02, 0·05 | **<0·001** | -0·005 | -0·02, 0·005 | 0·67 |
| Ab+ | 0·03 | 0·01, 0·06 | **0·01** | 0·04 | 0·02, 0·06 | **<0·001** | 0·06 | 0·04, 0·09 | **<0·001** | 0·006 | -0·010, 0·02 | 0·93 |
| **Model 3 - Fixed Effect[b]** | Est. | SE | P | Est. | SE | P | Est. | SE | P | Est. | SE | P |
| (Intercept) | 1·19 | 0·03 | **<0·001** | 1·38 | 0·04 | **<0·001** | 2·64 | 0·05 | **<0·001** | 0·19 | 0·03 | **<0·001** |
| Time | 0·007 | 0·009 | 0·46 | 0·01 | 0·007 | *0·05* | 0·03 | 0·008 | **0·001** | -0·002 | 0·006 | 0·72 |
| iMCI [iMCI] | 0·30 | 0·07 | **<0·001** | 0·14 | 0·09 | 0·13 | 0·40 | 0·12 | **<0·001** | -0·19 | 0·08 | *0·014* |
| Amyloid status [Ab+] | 0·05 | 0·04 | 0·14 | 0·06 | 0·05 | 0·23 | 0·09 | 0·06 | 0·15 | -0·0001 | 0·04 | 1·00 |
| Time:iMCI [iMCI] | 0·05 | 0·02 | **0·011** | -0·0008 | 0·02 | 0·96 | 0·02 | 0·02 | 0·24 | -0·01 | 0·01 | 0·26 |
| Time:Amyloid status [Ab+] | 0·01 | 0·01 | 0·33 | 0·03 | 0·01 | *0·014* | 0·03 | 0·01 | *0·013* | 0·01 | 0·008 | *0·08* |
| iMCI:Amyloid status [Ab+iMCI] | -0·26 | 0·09 | **0·005** | -0·01 | 0·13 | 0·93 | -0·20 | 0·16 | 0·21 | 0·28 | 0·11 | **0·010** |
| Time:iMCI:Amyloid status [Ab+iMCI] | 0·009 | 0·03 | 0·78 | 0·007 | 0·02 | 0·76 | 0·02 | 0·03 | 0·55 | -0·01 | 0·02 | 0·51 |
| **Slopes** | Est. | 95% CI | $P_{adj}$ | Est. | 95% CI | $P_{adj}$ | Est. | 95% CI | $P_{adj}$ | Est. | 95% CI | $P_{adj}$ |
| Ab-Stable | 0·008 | -0·01, 0·03 | 1·00 | 0·01 | 0·0007, 0·03 | 0·16 | 0·03 | 0·01, 0·04 | **0·002** | -0·003 | -0·01, 0·008 | 1·00 |
| Ab+Stable | 0·02 | -0·006, 0·05 | 0·52 | 0·04 | 0·02, 0·06 | **<0·001** | 0·06 | 0·03, 0·08 | **<0·001** | 0·01 | -0·005, 0·03 | 0·64 |
| Ab-iMCI | 0·06 | 0·02, 0·10 | **0·01** | 0·01 | -0·02, 0·04 | 1·00 | 0·05 | 0·01, 0·08 | **0·02** | -0·02 | -0·04, 0·007 | 0·64 |
| Ab+iMCI | 0·08 | 0·04, 0·13 | **0·001** | 0·05 | 0·01, 0·08 | **0·03** | 0·10 | 0·06, 0·14 | **<0·001** | -0·02 | -0·04, 0·01 | 1·00 |
| **Pairwise comparisons** | Est. | 95% CI | $P_{adj}$ | Slope | 95% CI | $P_{adj}$ | Slope | 95% CI | $P_{adj}$ | Slope | 95% CI | $P_{adj}$ |
| Ab-Stable / Ab+Stable | -0·01 | -0·04, 0·01 | 1·00 | -0·03 | -0·05, -0·005 | *0·08* | -0·03 | -0·05,-0·006 | *0·07* | -0·01 | -0·03, 0·001 | 0·44 |
| Ab-Stable / Ab-iMCI | -0·05 | -0·09, -0·01 | *0·06* | 0·0008 | -0·03, 0·03 | 1·00 | -0·02 | -0·06, 0·01 | 1·00 | 0·01 | -0·01, 0·04 | 1·00 |
| Ab-Stable / Ab+iMCI | -0·07 | -0·12, -0·03 | **0·006** | -0·03 | -0·07, 0·002 | 0·38 | -0·07 | -0·11, -0·03 | **0·006** | 0·01 | -0·02, 0·04 | 1·00 |
| Ab+Stable / Ab-iMCI | -0·04 | -0·08, 0·003 | 0·41 | 0·03 | -0·007, 0·06 | 0·72 | 0·008 | -0·03, 0·05 | 1·00 | 0·03 | 0·002, 0·06 | 0·21 |
| Ab+Stable / Ab+iMCI | -0·06 | -0·11, -0·01 | *0·06* | -0·007 | -0·04, 0·03 | 1·00 | -0·04 | -0·08, 0·004 | 0·47 | 0·03 | -0·003, 0·06 | 0·48 |
| Ab-iMCI / Ab+iMCI | -0·02 | -0·08, 0·03 | 1·00 | -0·03 | -0·07, 0·010 | 0·78 | -0·05 | -0·10, 0·005 | 0·45 | -0·002 | -0·04, 0·03 | 1·00 |

To reduce the number of lines, those referring to covariates are not presented above. [a] Adjusted for age, sex, years of education, cohort (only for combined analyses), setting (only for DELCODE), and their interaction with time. [b] Adjusted in addition for amyloid modality (PET, CSF, plasma).

**eTable 6.** Detailed results from Model 1 according to amyloid status subgroups.

| | **Model 1** | **SP-Ecog** | | | **Self-Ecog** | | | **Cum-Ecog** | | | **CAI-Ecog** | | |
|---|---|---|---|---|---|---|---|---|---|---|---|---|---|
| **Combined sample** | **Ab- participants - Fixed Effect** | Est. | SE | P | Est. | SE | P | Est. | SE | P | Est. | SE | P |
| | (Intercept) | 1·19 | 0·02 | **<0·001** | 1·22 | 0·03 | **<0·001** | 2·40 | 0·04 | **<0·001** | 0·01 | 0·03 | 0·67 |
| | Time | 0·007 | 0·005 | 0·11 | 0·01 | 0·005 | *0·02* | 0·02 | 0·007 | **0·004** | 0·006 | 0·005 | 0·20 |
| | iMCI [iMCI] | 0·17 | 0·03 | **<0·001** | 0·07 | 0·04 | *0·09* | 0·24 | 0·05 | **<0·001** | -0·09 | 0·04 | *0·03* |
| | Time:iMCI [iMCI] | 0·01 | 0·007 | *0·04* | 0·01 | 0·007 | *0·06* | 0·02 | 0·01 | *0·02* | 0·0008 | 0·007 | 0·90 |
| | **Ab+ participants - Fixed Effect** | Est. | SE | P | Est. | SE | P | Est. | SE | P | Est. | SE | P |
| | (Intercept) | 1·15 | 0·04 | **<0·001** | 1·22 | 0·05 | **<0·001** | 2·35 | 0·07 | **<0·001** | 0·06 | 0·06 | 0·32 |
| | Time | 0·02 | 0·009 | *0·03* | 0·02 | 0·01 | 0·04 | 0·04 | 0·01 | **0·003** | 0·005 | 0·01 | 0·61 |
| | iMCI [iMCI] | 0·07 | 0·04 | *0·09* | 0·09 | 0·05 | *0·08* | 0·17 | 0·07 | *0·02* | 0·02 | 0·06 | 0·77 |
| | Time:iMCI [iMCI] | 0·07 | 0·01 | **<0·001** | 0·02 | 0·01 | *0·08* | 0·08 | 0·02 | **<0·001** | -0·03 | 0·01 | **0·003** |
| **DELCODE** | **Ab- participants - Fixed Effect** | Est. | SE | P | Est. | SE | P | Est. | SE | P | Est. | SE | P |
| | (Intercept) | 1·22 | 0·03 | **<0·001** | 1·22 | 0·03 | **<0·001** | 2·42 | 0·04 | **<0·001** | -0·009 | 0·04 | 0·82 |
| | Time | 0·006 | 0·004 | 0·13 | 0·01 | 0·005 | **0·009** | 0·02 | 0·007 | **0·002** | 0·009 | 0·006 | 0·11 |
| | iMCI [iMCI] | 0·12 | 0·04 | **0·001** | 0·04 | 0·04 | 0·32 | 0·19 | 0·06 | **0·002** | -0·06 | 0·05 | 0·30 |
| | Time:iMCI [iMCI] | 0·008 | 0·007 | 0·23 | 0·02 | 0·008 | *0·02* | 0·02 | 0·01 | *0·04* | 0·005 | 0·01 | 0·58 |
| | **Ab+ participants - Fixed Effect** | Est. | SE | P | Est. | SE | P | Est. | SE | P | Est. | SE | P |
| | (Intercept) | 1·15 | 0·05 | **<0·001** | 1·22 | 0·05 | **<0·001** | 2·36 | 0·07 | **<0·001** | 0·06 | 0·06 | 0·33 |
| | Time | 0·02 | 0·009 | *0·02* | 0·02 | 0·01 | 0·13 | 0·04 | 0·02 | *0·02* | -0·0002 | 0·01 | 0·99 |
| | iMCI [iMCI] | 0·10 | 0·06 | *0·07* | 0·07 | 0·07 | 0·31 | 0·16 | 0·09 | *0·08* | -0·04 | 0·08 | 0·56 |
| | Time:iMCI [iMCI] | 0·07 | 0·01 | **<0·001** | 0·03 | 0·01 | *0·015* | 0·11 | 0·02 | **<0·001** | -0·03 | 0·01 | *0·03* |
| **ADNI** | **Ab- participants - Fixed Effect** | Est. | SE | P | Est. | SE | P | Est. | SE | P | Est. | SE | P |
| | (Intercept) | 1·18 | 0·03 | **<0·001** | 1·37 | 0·04 | **<0·001** | 2·63 | 0·05 | **<0·001** | 0·18 | 0·03 | **<0·001** |
| | Time | 0·009 | 0·01 | 0·38 | 0·009 | 0·007 | 0·17 | 0·02 | 0·009 | **0·009** | -0·006 | 0·005 | 0·23 |
| | iMCI [iMCI] | 0·31 | 0·07 | **<0·001** | 0·14 | 0·09 | 0·13 | 0·40 | 0·12 | **<0·001** | -0·21 | 0·07 | **0·003** |
| | Time:iMCI [iMCI] | 0·05 | 0·02 | *0·013* | 0·0006 | 0·01 | 0·97 | 0·03 | 0·02 | 0·16 | -0·009 | 0·010 | 0·36 |
| | **Ab+ participants - Fixed Effect** | Est. | SE | P | Est. | SE | P | Est. | SE | P | Est. | SE | P |
| | (Intercept) | 1·23 | 0·05 | **<0·001** | 1·43 | 0·07 | **<0·001** | 2·71 | 0·09 | **<0·001** | 0·18 | 0·07 | **0·010** |
| | Time | 0·03 | 0·02 | *0·08* | 0·06 | 0·02 | **0·002** | 0·08 | 0·02 | **<0·001** | 0·02 | 0·01 | 0·14 |
| | iMCI [iMCI] | 0·03 | 0·07 | 0·64 | 0·17 | 0·10 | 0·10 | 0·21 | 0·13 | 0·10 | 0·13 | 0·09 | 0·18 |
| | Time:iMCI [iMCI] | 0·07 | 0·03 | **0·012** | -0·007 | 0·02 | 0·77 | 0·04 | 0·03 | 0·17 | -0·04 | 0·02 | *0·05* |

To reduce the number of lines, those referring to covariates are not presented above. Analyses are all adjusted for age, sex, years of education, cohort (only for combined analyses), setting (only for DELCODE), and their interaction with time.

**eTable 7.** Results from Models 1-3 restricted to SCD patients from the DELCODE cohort, including subgroup slopes and pairwise comparisons with Bonferroni-adjusted p-values using the ggeffects::hypothesis_test() function.

| | **SP-Ecog** | | | **Self-Ecog** | | | **Cum-Ecog** | | | **CAI-Ecog** | | |
|---|---|---|---|---|---|---|---|---|---|---|---|---|
| **Model 1 - Fixed Effect[a]** | Est. | SE | P | Est. | SE | P | Est. | SE | P | Est. | SE | P |
| (Intercept) | 1·41 | 0·03 | **<0·001** | 1·53 | 0·04 | **<0·001** | 2·92 | 0·05 | **<0·001** | 0·09 | 0·04 | *0·04* |
| Time | 0·02 | 0·006 | **0·002** | 0·01 | 0·007 | 0·11 | 0·04 | 0·01 | **0·002** | -0·003 | 0·008 | 0·69 |
| iMCI [iMCI] | 0·09 | 0·04 | *0·05* | 0·03 | 0·05 | 0·52 | 0·13 | 0·07 | *0·06* | -0·04 | 0·06 | 0·53 |
| Time:iMCI [iMCI] | 0·05 | 0·01 | **<0·001** | 0·03 | 0·01 | **0·003** | 0·08 | 0·02 | **<0·001** | -0·02 | 0·01 | 0·19 |
| **Slopes** | Est. | 95% CI | $P_{adj}$ | Est. | 95% CI | $P_{adj}$ | Est. | 95% CI | $P_{adj}$ | Est. | 95% CI | $P_{adj}$ |
| Stable | 0·02 | 0·009, 0·03 | **<0·001** | 0·01 | 0·0005, 0·03 | *0·08* | 0·04 | 0·02, 0·06 | **<0·001** | -0·002 | -0·02, 0·01 | 1·00 |
| iMCI | 0·07 | 0·05, 0·09 | **<0·001** | 0·05 | 0·03, 0·07 | **<0·001** | 0·12 | 0·09, 0·15 | **<0·001** | -0·02 | -0·04, 0·004 | 0·22 |
| **Model 2 - Fixed Effect[b]** | Est. | SE | P | Est. | SE | P | Est. | SE | P | Est. | SE | P |
| (Intercept) | 1·45 | 0·03 | **<0·001** | 1·56 | 0·04 | **<0·001** | 2·98 | 0·06 | **<0·001** | 0·09 | 0·05 | *0·08* |
| Time | 0·02 | 0·007 | **0·01** | 0·01 | 0·008 | 0·21 | 0·04 | 0·01 | **0·008** | 0·001 | 0·009 | 0·89 |
| Amyloid status [Ab+] | -0·02 | 0·04 | 0·55 | 0·001 | 0·05 | 0·98 | -0·03 | 0·07 | 0·70 | 0·02 | 0·06 | 0·77 |
| Time:Amyloid status [Ab+] | 0·05 | 0·009 | **<0·001** | 0·02 | 0·01 | *0·03* | 0·07 | 0·02 | **<0·001** | -0·03 | 0·01 | **0·006** |
| **Slopes** | Est. | 95% CI | $P_{adj}$ | Est. | 95% CI | $P_{adj}$ | Est. | 95% CI | $P_{adj}$ | Est. | 95% CI | $P_{adj}$ |
| Ab- | 0·02 | 0·006, 0·04 | **0·009** | 0·01 | -0·003, 0·03 | 0·22 | 0·04 | 0·01, 0·07 | **0·005** | 0·002 | -0·02, 0·02 | 1·00 |
| Ab+ | 0·07 | 0·05, 0·09 | **<0·001** | 0·03 | 0·02, 0·05 | **<0·001** | 0·11 | 0·08, 0·14 | **<0·001** | -0·03 | -0·05, -0·008 | **0·01** |
| **Model 3 - Fixed Effect[b]** | Est. | SE | P | Est. | SE | P | Est. | SE | P | Est. | SE | P |
| (Intercept) | 1·42 | 0·04 | **<0·001** | 1·55 | 0·05 | **<0·001** | 2·94 | 0·06 | **<0·001** | 0·10 | 0·06 | *0·07* |
| Time | 0·02 | 0·007 | *0·04* | 0·005 | 0·009 | 0·58 | 0·03 | 0·01 | *0·05* | -0·0001 | 0·01 | 0·99 |
| iMCI [iMCI] | 0·11 | 0·06 | *0·07* | 0·04 | 0·07 | 0·58 | 0·17 | 0·09 | *0·07* | -0·04 | 0·09 | 0·66 |
| Amyloid status [Ab+] | -0·03 | 0·05 | 0·55 | 0·006 | 0·06 | 0·92 | -0·02 | 0·08 | 0·81 | 0·04 | 0·07 | 0·63 |
| Time:iMCI [iMCI] | 0·01 | 0·01 | 0·41 | 0·02 | 0·01 | *0·09* | 0·03 | 0·02 | 0·15 | 0·01 | 0·02 | 0·55 |
| Time:Amyloid status [Ab+] | 0·03 | 0·009 | **0·008** | 0·01 | 0·01 | 0·21 | 0·04 | 0·02 | *0·04* | -0·02 | 0·01 | 0·18 |
| iMCI:Amyloid status [Ab+iMCI] | -0·03 | 0·09 | 0·74 | -0·03 | 0·11 | 0·81 | -0·08 | 0·14 | 0·56 | -0·03 | 0·13 | 0·80 |
| Time:iMCI:Amyloid status [Ab+iMCI] | 0·06 | 0·02 | **<0·001** | 0·01 | 0·02 | 0·61 | 0·09 | 0·03 | **0·078** | -0·04 | 0·02 | *0·08* |
| **Slopes** | Est. | 95% CI | $P_{adj}$ | Est. | 95% CI | $P_{adj}$ | Est. | 95% CI | $P_{adj}$ | Est. | 95% CI | $P_{adj}$ |
| Ab-Stable | 0·02 | 0·002, 0·03 | *0·09* | 0·007 | -0·01, 0·02 | 1·00 | 0·03 | 0·004, 0·06 | *0·10* | -0·0009 | -0·02, 0·02 | 1·00 |
| Ab+Stable | 0·04 | 0·02, 0·06 | **<0·001** | 0·02 | 0·0005, 0·04 | 0·18 | 0·07 | 0·03, 0·10 | **<0·001** | -0·02 | -0·04, 0·008 | 0·73 |
| Ab-iMCI | 0·03 | 0·003, 0·05 | 0·10 | 0·03 | 0·004, 0·06 | 0·09 | 0·06 | 0·02, 0·10 | **0·01** | 0·01 | -0·02, 0·04 | 1·00 |
| Ab+iMCI | 0·12 | 0·09, 0·14 | **<0·001** | 0·06 | 0·03, 0·08 | **<0·001** | 0·18 | 0·14, 0·22 | **<0·001** | -0·05 | -0·08, -0·02 | **0·006** |

| | SP-Ecog | | | Self-Ecog | | | Cum-Ecog | | | CAI-Ecog | | |
|---|---|---|---|---|---|---|---|---|---|---|---|---|
| **Pairwise comparisons** | Est. | 95% CI | $P_{adj}$ | Slope | 95% CI | $P_{adj}$ | Slope | 95% CI | $P_{adj}$ | Slope | 95% CI | $P_{adj}$ |
| Ab-Stable / Ab+Stable | -0·03 | -0·04, -0·007 | **0·04** | -0·01 | -0·04, 0·008 | 1·00 | -0·04 | -0·07,-0·002 | 0·22 | 0·02 | -0·008, 0·04 | 1·00 |
| Ab-Stable / Ab-iMCI | -0·01 | -0·03, 0·01 | 1·00 | -0·02 | -0·05, 0·004 | 0·54 | -0·03 | -0·07, 0·01 | 0·87 | -0·01 | -0·04, 0·02 | 1·00 |
| Ab-Stable / Ab+iMCI | -0·10 | -0·12, -0·08 | **<0·001** | -0·05 | -0·08, -0·02 | **0·003** | -0·15 | -0·20, -0·11 | **<0·001** | 0·05 | 0·02, 0·08 | **0·02** |
| Ab+Stable / Ab-iMCI | 0·02 | -0·, 0·04 | 1·00 | -0·01 | -0·04, 0·02 | 1·00 | 0·004 | -0·04, 0·05 | 1·00 | -0·03 | -0·06, 0·009 | 0·85 |
| Ab+Stable / Ab+iMCI | -0·07 | -0·10, -0·05 | **<0·001** | -0·04 | -0·07, -0·005 | 0·13 | -0·12 | -0·16, -0·07 | **<0·001** | 0·03 | -0·002, 0·07 | 0·41 |
| Ab-iMCI / Ab+iMCI | -0·09 | -0·12, -0·06 | **<0·001** | -0·03 | -0·06, 0·01 | 0·97 | -0·12 | -0·18, -0·07 | **<0·001** | 0·06 | 0·02, 0·10 | **0·02** |
| **Model 1 in Ab- SCD patients** | Est. | SE | P | Est. | SE | P | Est. | SE | P | Est. | SE | P |
| (Intercept) | 1·44 | 0·03 | **<0·001** | 1·53 | 0·04 | **<0·001** | 2·94 | 0·06 | **<0·001** | 0·07 | 0·05 | 0·21 |
| Time | 0·003 | 0·007 | 0·67 | 0·008 | 0·008 | 0·32 | 0·02 | 0·01 | 0·13 | 0·009 | 0·009 | 0·35 |
| iMCI [iMCI] | 0·11 | 0·06 | *0·06* | 0·05 | 0·07 | 0·42 | 0·19 | 0·09 | *0·04* | -0·02 | 0·09 | 0·79 |
| Time:iMCI [iMCI] | 0·01 | 0·01 | 0·18 | 0·03 | 0·01 | *0·03* | 0·04 | 0·02 | *0·05* | 0·008 | 0·02 | 0·59 |
| **Model 1 in Ab+ SCD patients** | Est. | SE | P | Est. | SE | P | Est. | SE | P | Est. | SE | P |
| (Intercept) | 1·35 | 0·05 | **<0·001** | 1·50 | 0·07 | **<0·001** | 2·83 | 0·09 | **<0·001** | 0·13 | 0·08 | 0·12 |
| Time | 0·03 | 0·01 | **0·003** | 0·008 | 0·01 | 0·56 | 0·04 | 0·02 | *0·07* | -0·02 | 0·01 | 0·16 |
| iMCI [iMCI] | 0·07 | 0·07 | 0·32 | 0·009 | 0·09 | 0·92 | 0·08 | 0·12 | 0·50 | -0·07 | 0·10 | 0·53 |
| Time:iMCI [iMCI] | 0·07 | 0·02 | **<0·001** | 0·03 | 0·02 | 0·10 | 0·11 | 0·03 | **<0·001** | -0·04 | 0·02 | *0·05* |

To reduce the number of lines, those referring to covariates are not presented above. [a] Adjusted for age, sex, years of education, and their interaction with time. [b] Adjusted in addition for amyloid modality (PET, CSF, plasma).

**eTable 8.** Risk of clinical progression to incident mild cognitive impairment (iMCI) based on one-year changes in Ecog scores, stratified by amyloid status.

**eTable 6A**. Combined sample.

| | | **SP-Ecog** | | | **Self-Ecog** | | | **Cum-Ecog** | | | **CAI-Ecog** | |
|---|---|---|---|---|---|---|---|---|---|---|---|---|
| **iMCI** | $N_{obs[event]}$ | HR [95% CI] | P | $N_{obs[event]}$ | HR [95% CI] | P | $N_{obs[event]}$ | HR [95% CI] | P | $N_{obs[event]}$ | HR [95% CI] | P |
| Ecog scores | | | **0·002** | | | 0·13 | | | *0·019* | | | 0·59 |
| Unchanged | 243 [26] | | | 222 [27] | | | 247 [27] | | | 255 [32] | | |
| Decrease | 47 [6] | 1·35 [0·50, 3·61] | 0·55 | 64 [11] | 0·99 [0·45, 2·17] | 0·98 | 48 [8] | 1·03 [0·44, 2·42] | 0·95 | 54 [14] | 1·44 [0·72, 2·89] | 0·30 |
| Increase | 63 [20] | 3·24 [1·73, 6·07] | **<0·001** | 67 [14] | 1·97 [1·01, 3·86] | *0·048* | 58 [17] | 2·54 [1·31, 4·91] | **0·005** | 44 [6] | 1·02 [0·41, 2·53] | 0·97 |
| Baseline levels | 353 [52] | 1·11 [0·43, 2·85] | 0·83 | 353 [52] | 1·29 [0·50, 3·35] | 0·59 | 353 [52] | 1·23 [0·70, 2·15] | 0·47 | 353 [52] | 0·83 [0·39, 1·76] | 0·63 |
| **iMCI*Ab** | $N_{obs[event]}$ | HR [95% CI] | P | $N_{obs[event]}$ | HR [95% CI] | P | $N_{obs[event]}$ | HR [95% CI] | P | $N_{obs[event]}$ | HR [95% CI] | P |
| Ecog scores | | | 0·20 | | | 0·90 | | | **0·002** | | | 0·50 |
| Unchanged | 243 [26] | | | 222 [27] | | | 247 [27] | | | 255 [32] | | |
| Decrease | 47 [6] | 2·07 [0·61, 7·05] | 0·80 | 64 [11] | 1·11 [0·39, 3·14] | 0·80 | 48 [8] | 1·61 [0·50, 5·16] | 0·40 | 54 [14] | 1·75 [0·70, 4·37] | 0·20 |
| Increase | 63 [20] | 2·42 [0·89, 6·57] | *0·08* | 67 [14] | 1·32 [0·47, 3·74] | 0·60 | 58 [17] | 6·04 [2·27, 16·1] | **<0·001** | 44 [6] | 1·32 [0·47, 3·74] | 0·90 |
| Baseline levels | 353 [52] | 1·64 [0·46, 5·79] | 0·40 | 353 [52] | 1·43 [0·45, 4·53] | 0·50 | 353 [52] | 1·24 [0·60, 2·54] | 0·60 | 353 [52] | 1·43 [0·45, 4·53] | 0·20 |
| Ecog scores * Amyloid status | | | 0·50 | | | 0·30 | | | *0·09* | | | 0·80 |
| Decrease * Aβ+ | 18 [2] | 0·45 [0·05, 3·80] | 0·50 | 21 [4] | 0·98 [0·19, 5·11] | >0·90 | 15 [3] | 0·45 [0·08, 2·72] | 0·40 | 17 [6] | 0·82 [0·20, 3·35] | 0·80 |
| Increase * Aβ+ | 26 [12] | 1·64 [0·43, 6·22] | 0·50 | 27 [9] | 2·76 [0·70, 10·90] | 0·15 | 25 [7] | 0·22 [0·06, 0·87] | *0·03* | 20 [4] | 1·60 [0·24, 10·50] | 0·60 |
| Baseline levels * Amyloid status | 123 [26] | 0·43 [0·06, 3·05] | 0·40 | 123 [26] | 0·77 [0·10, 5·80] | 0·80 | 123 [26] | 0·77 [0·26, 2·32] | 0·60 | 123 [26] | 3·08 [0·68, 13·80] | 0·14 |
| **iMCI in Ab-** | $N_{obs[event]}$ | HR [95% CI] | P | $N_{obs[event]}$ | HR [95% CI] | P | $N_{obs[event]}$ | HR [95% CI] | P | $N_{obs[event]}$ | HR [95% CI] | P |
| Ecog scores | | | 0·13 | | | 0·83 | | | *0·014* | | | 0·62 |
| Unchanged | 164 [14] | | | 147 [14] | | | 164 [11] | | | 169 [16] | | |
| Decrease | 29 [4] | 2·55 [0·72, 9·03] | 0·15 | 43 [7] | 1·17 [0·39, 3·50] | 0·78 | 33 [5] | 1·44 [0·40, 5·15] | 0·58 | 37 [8] | 1·51 [0·57, 3·96] | 0·41 |
| Increase | 37 [8] | 2·83 [0·99, 8·07] | *0·05* | 40 [5] | 1·40 [0·49, 4·01] | 0·54 | 33 [10] | 4·73 [1·66, 13·5] | **0·004** | 24 [2] | 0·77 [0·17, 3·59] | 0·74 |
| Baseline levels | 230 [26] | 1·70 [0·43, 6·78] | 0·45 | 230 [26] | 1·26 [0·36, 4·40] | 0·72 | 230 [26] | 1·25 [0·55, 2·84] | 0·59 | 230 [26] | 0·59 [0·21, 1·68] | 0·33 |
| **iMCI in Ab+** | $N_{obs[event]}$ | HR [95% CI] | P | $N_{obs[event]}$ | HR [95% CI] | P | $N_{obs[event]}$ | HR [95% CI] | P | $N_{obs[event]}$ | HR [95% CI] | P |
| Ecog scores | | | **0·003** | | | 0·11 | | | 0·57 | | | 0·82 |
| Unchanged | 79 [12] | | | 75 [13] | | | 83 [16] | | | 86 [16] | | |
| Decrease | 18 [2] | 0·60 [0·11, 3·22] | 0·55 | 21 [4] | 0·75 [0·21, 2·72] | 0·66 | 15 [3] | 0·63 [0·16, 2·41] | 0·49 | 17 [6] | 1·42 [0·46, 4·42] | 0·54 |
| Increase | 26 [12] | 4·18 [1·80, 9·73] | **<0·001** | 27 [9] | 2·44 [0·92, 6·46] | *0·07* | 25 [7] | 1·37 [0·52, 3·62] | 0·53 | 20 [4] | 0·98 [0·31, 3·10] | 0·97 |
| Baseline levels | 123 [26] | 1·41 [0·26, 7·73] | 0·69 | 123 [26] | 1·89 [0·35, 10·3] | 0·46 | 123 [26] | 1·32 [0·49, 3·55] | 0·58 | 123 [26] | 1·29 [0·40, 4·15] | 0·66 |

**eTable 6B**· DELCODE cohort·

| **DELCODE** | | **SP-Ecog** | | | **Self-Ecog** | | | **Cum-Ecog** | | | **CAI-Ecog** | |
|---|---|---|---|---|---|---|---|---|---|---|---|---|
| **iMCI** | $N_{obs[event]}$ | HR [95% CI] | P | $N_{obs[event]}$ | HR [95% CI] | P | $N_{obs[event]}$ | HR [95% CI] | P | $N_{obs[event]}$ | HR [95% CI] | P |
| Overall | | | **0·008** | | | 0·15 | | | **0·007** | | | 0·54 |
| Unchanged | 148 [20] | | | 143 [21] | | | 154 [19] | | | 165 [26] | | |
| Decrease | 30 [5] | 1·72 [0·45, 3·14] | 0·33 | 40 [7] | 0·99 [0·38, 2·61] | 0·99 | 30 [7] | 1·89 [0·72, 4·94] | 0·19 | 33 [9] | 1·51 [0·67, 3·40] | 0·32 |
| Increase | 37 [13] | 3·61 [1·69, 5·94] | **<0·001** | 32 [10] | 2·18 [1·01, 4·70] | *0·047* | 31 [12] | 3·70 [1·69, 8·08] | **0·001** | 17 [3] | 0·82 [0·24, 2·82] | 0·74 |
| Baseline levels | 215 [38] | 0·70 [0·46, 2·99] | 0·52 | 215 [38] | 1·12 [0·35, 3·60] | 0·85 | 215 [38] | 0·87 [0·44, 1·72] | 0·68 | 215 [38] | 0·91 [0·39, 2·11] | 0·83 |
| **iMCI in Ab-** | $N_{obs[event]}$ | HR [95% CI] | P | $N_{obs[event]}$ | HR [95% CI] | P | $N_{obs[event]}$ | HR [95% CI] | P | $N_{obs[event]}$ | HR [95% CI] | P |
| Overall | | | *0·07* | | | 0·74 | | | **0·007** | | | 0·91 |
| Unchanged | 102 [10] | | | 97 [12] | | | 102 [8] | | | 109 [13] | | |
| Decrease | 19 [4] | 3·92 [0·56, 6·75] | *0·05* | 29 [4] | 0·97 [0·27, 3·49] | 0·96 | 23 [4] | 2·66 [0·68, 10·4] | 0·16 | 25 [5] | 1·28 [0·40, 4·05] | 0·68 |
| Increase | 22 [6] | 3·45 [0·92, 7·51] | *0·05* | 17 [4] | 1·57 [0·50, 4·98] | 0·44 | 18 [8] | 7·56 [2·20, 26·0] | **0·001** | 9 [2] | 0·98 [0·20, 4·90] | 0·98 |
| Baseline levels | 143 [20] | 0·71 [0·48, 7·35] | 0·69 | 143 [20] | 1·36 [0·33, 5·52] | 0·67 | 143 [20] | 0·89 [0·33, 2·38] | 0·81 | 143 [20] | 0·88 [0·27, 2·87] | 0·84 |
| **iMCI in Ab+** | $N_{obs[event]}$ | HR [95% CI] | P | $N_{obs[event]}$ | HR [95% CI] | P | $N_{obs[event]}$ | HR [95% CI] | P | $N_{obs[event]}$ | HR [95% CI] | P |
| Overall | | | *0·03* | | | 0·21 | | | 0·56 | | | 0·21 |
| Unchanged | 46 [10] | | | 46 [9] | | | 52 [11] | | | 56 [13] | | |
| Decrease | 11 [1] | 0·43 [0·05, 4·10] | 0·46 | 11 [3] | 0·84 [0·21, 5·07] | 0·97 | 7 [3] | 1·95 [0·44, 8·56] | 0·38 | 8 [4] | 2·69 [0·81, 8·90] | 0·11 |
| Increase | 15 [7] | 3·77 [1·28, 11·1] | *0·02* | 15 [6] | 1·79 [0·87, 9·70] | *0·08* | 13 [4] | 1·61 [0·50, 5·22] | 0·43 | 8 [1] | 0·48 [0·06, 3·74] | 0·49 |
| Baseline levels | 72 [18] | 1·20 [0·14, 10·4] | 0·87 | 72 [18] | 1·48 [0·16, 11·7] | 0·79 | 72 [18] | 0·87 [0·24, 3·24] | 0·84 | 72 [18] | 1·07 [0·28, 4·04] | 0·92 |
| **DELCODE, SCD patients only** | | **SP-Ecog** | | | **Self-Ecog** | | | **Cum-Ecog** | | | **CAI-Ecog** | |
| **iMCI** | $N_{obs[event]}$ | HR [95% CI] | P | $N_{obs[event]}$ | HR [95% CI] | P | $N_{obs[event]}$ | HR [95% CI] | P | $N_{obs[event]}$ | HR [95% CI] | P |
| Overall | | | 0·19 | | | 0·13 | | | 0·26 | | | 0·89 |
| Unchanged | 90 [16] | | | 85 [16] | | | 94 [16] | | | 94 [16] | | |
| Decrease | 24 [4] | 1·59 [0·48, 5·29] | 0·45 | 31 [4] | 0·74 [0·22, 2·47] | 0·62 | 24 [4] | 1·04 [0·32, 3·35] | 0·95 | 24 [4] | 1·51 [0·67, 3·40] | 0·90 |
| Increase | 29 [9] | 2·40 [0·96, 5·98] | *0·06* | 27 [9] | 2·22 [0·96, 5·13] | *0·06* | 25 [9] | 2·08 [0·88, 4·94] | 0·10 | 25 [9] | 0·82 [0·24, 2·82] | 0·64 |
| Baseline levels | 143 [29] | 0·37 [0·11, 1·29] | 0·12 | 143 [29] | 0·69 [0·19, 2·56] | 0·58 | 143 [29] | 0·61 [0·29, 1·27] | 0·18 | 143 [29] | 0·91 [0·39, 2·11] | 0·70 |
| **iMCI in Ab-** | $N_{obs[event]}$ | HR [95% CI] | P | $N_{obs[event]}$ | HR [95% CI] | P | $N_{obs[event]}$ | HR [95% CI] | P | $N_{obs[event]}$ | HR [95% CI] | P |
| Overall | | | 0·29 | | | 0·43 | | | 0·22 | | | 0·43 |
| Unchanged | 61 [8] | | | 58 [8] | | | 60 [6] | | | 67 [10] | | |
| Decrease | 16 [3] | 3·66 [0·77,17·3] | 0·10 | 24 [3] | 1·12 [0·25, 5·02] | 0·88 | 20 [3] | 1·70 [0·38, 7·60] | 0·49 | 20 [3] | 0·40 [0·09, 1·75] | 0·22 |
| Increase | 19 [4] | 1·76 [0·41, 7·60] | 0·45 | 14 [4] | 2·39 [0·67, 8·45] | 0·18 | 16 [6] | 3·49 [0·88, 13·9] | *0·07* | 9 [2] | 0·60 [0·12, 3·03] | 0·54 |
| Baseline levels | 96 [15] | 0·19 [0·03, 1·51] | 0·12 | 96 [15] | 0·88 [0·19, 4·10] | 0·87 | 96 [15] | 0·51 [0·17, 1·54] | 0·23 | 96 [15] | 1·95 [0·50, 7·58] | 0·34 |
| **iMCI in Ab+** | $N_{obs[event]}$ | HR [95% CI] | P | $N_{obs[event]}$ | HR [95% CI] | P | $N_{obs[event]}$ | HR [95% CI] | P | $N_{obs[event]}$ | HR [95% CI] | P |
| Overall | | | *0·09* | | | *0·44* | | | 0·97 | | | 0·26 |
| Unchanged | 29 [8] | | | 27 [8] | | | 34 [10] | | | 36 [10] | | |
| Decrease | 8 [1] | 0·59 [0·06, 5·75] | 0·65 | 7 [1] | 0·59 [0·05, 5·37] | 0·59 | 4 [1] | 0·84 [0·09, 7·93] | 0·88 | 5 [3] | 3·10 [0·73, 13·1] | 0·12 |
| Increase | 10 [5] | 3·95 [1·11, 14·1] | *0·03* | 13 [5] | 3·95 [0·51, 6·71] | *0·35* | 9 [3] | 1·15 [0·30, 4·43] | 0·83 | 6 [1] | 0·54 [0·07, 4·32] | 0·57 |
| Baseline levels | 47 [14] | 0·71 [0·08, 6·55] | 0·76 | 47 [14] | 0·71 [0·05, 7·19] | 0·69 | 47 [14] | 0·54 [0·12, 2·50] | 0·43 | 47 [14] | 1·05 [0·25, 4·43] | 0·94 |

**eTable 6C**· ADNI cohort·

| | SP-Ecog | | | Self-Ecog | | | Cum-Ecog | | | CAI-Ecog | | |
|---|---|---|---|---|---|---|---|---|---|---|---|---|
| **iMCI** | $N_{obs[event]}$ | HR [95% CI] | P | $N_{obs[event]}$ | HR [95% CI] | P | $N_{obs[event]}$ | HR [95% CI] | P | $N_{obs[event]}$ | HR [95% CI] | P |
| Overall | | | 0·13 | | | 0·85 | | | *0·08* | | | 0·97 |
| Unchanged | 95 [6] | | | 79 [6] | | | 93 [8] | | | 90 [6] | | |
| Decrease | 17 [1] | 0·49 [0·05, 4·52] | 0·53 | 24 [4] | 0·93 [0·20, 4·26] | 0·93 | 18 [1] | 0·13 [0·01, 1·26] | *0·08* | 21 [5] | 1·08 [0·21, 5·57] | 0·92 |
| Increase | 26 [7] | 3·12 [0·88, 11·1] | *0·08* | 35 [4] | 1·43 [0·33, 6·22] | 0·63 | 27 [5] | 1·01 [0·29, 3·56] | 0·98 | 27 [3] | 1·21 [0·29, 5·08] | 0·79 |
| Baseline levels | 138 [14] | 6·61 [1·50, 29·1] | **0·012** | 138 [14] | 2·18 [0·38, 12·6] | 0·38 | 138 [14] | 2·97 [1·26, 7·02] | *0·013* | 138 [14] | 0·41 [0·06, 2·94] | 0·38 |
| **iMCI in Ab-** | $N_{obs[event]}$ | HR [95% CI] | P | $N_{obs[event]}$ | HR [95% CI] | P | $N_{obs[event]}$ | HR [95% CI] | P | $N_{obs[event]}$ | HR [95% CI] | P |
| Overall | | | 0·24 | | | 0·83 | | | 0·12 | | | 0·52 |
| Unchanged | 62 [4] | | | 50 [2] | | | 62 [3] | | | 60 [3] | | |
| Decrease | 10 [0] | 0·00 [0·00, Inf] | 1·00 | 14 [3] | 2·01 [0·16, 25·2] | 0·59 | 10 [1] | 0·00 [0·00, Inf] | 1·00 | 12 [3] | 1·97 [0·20, 19·4] | 0·56 |
| Increase | 15 [2] | 4·61 [0·48, 44·8] | 0·19 | 23 [1] | 1·03 [0·06, 17·1] | 0·98 | 15 [2] | 1·64 [0·23, 11·8] | 0·62 | 15 [0] | 0·00 [0·00, Inf] | 1·00 |
| Baseline levels | 87 [6] | 16·58 [1·61,171] | *0·02* | 87 [6] | 1·56 [0·08, 31·8] | 0·77 | 87 [6] | 4·06 [1·08, 15·2] | *0·04* | 87 [6] | 0·07 [0·00, 1·62] | 0·10 |
| **iMCI in Ab+** | $N_{obs[event]}$ | HR [95% CI] | P | $N_{obs[event]}$ | HR [95% CI] | P | $N_{obs[event]}$ | HR [95% CI] | P | $N_{obs[event]}$ | HR [95% CI] | P |
| Overall | | | 0·13 | | | 0·25 | | | *0·02* | | | 0·23 |
| Unchanged | 33 [2] | | | 29 [4] | | | 31 [5] | | | 30 [3] | | |
| Decrease | 7 [1] | 1·26 [0·04, 38·8] | 0·89 | 10 [1] | 0·20 [0·01, 2·84] | 0·23 | 8 [0] | 0·00 [0·00, Inf] | 1·00 | 9 [2] | 0·08 [0·00, 2·24] | 0·14 |
| Increase | 11 [5] | 7·22 [0·66, 79·3] | 0·11 | 12 [3] | 1·64 [0·25, 10·8] | 0·61 | 12 [3] | 1·13 [0·14, 9·16] | 0·91 | 12 [3] | 0·65 [0·10, 4·16] | 0·65 |
| Baseline levels | 51 [8] | 1·24 [0·01, 210] | 0·93 | 51 [8] | 7·03 [0·34, 146] | 0·21 | 51 [8] | 4·58 [0·46, 45·7] | 0·20 | 51 [8] | 3·29 [0·25, 43·4] | 0·37 |

**eTable 9.** Risk of clinical progression to incident mild cognitive impairment (iMCI) based on one-year changes in Ecog scores, joint model including self- and study partner-reports.

| | | **Combined sample** | | | **DELCODE** | | | **ADNI** | | | **DELCODE, SCD patients** | |
|---|---|---|---|---|---|---|---|---|---|---|---|---|
| **iMCI** | $N_{obs[event]}$ | HR [95% CI] | P | $N_{obs[event]}$ | HR [95% CI] | P | $N_{obs[event]}$ | HR [95% CI] | P | $N_{obs[event]}$ | HR [95% CI] | P |
| SP-Ecog scores | | | **0·002** | | | **0·008** | | | 0·13 | | | **0·008** |
| Unchanged | 243 [26] | | | 148 [20] | | | 95 [6] | | | 148 [20] | | |
| Decrease | 47 [6] | 1·45 [0·54, 3·90] | 0·50 | 30 [5] | 2·12 [0·70, 6·38] | 0·20 | 17 [1] | 0·43 [0·04, 4·62] | 0·50 | 30 [5] | 1·72 [0·45, 3·14] | 0·33 |
| Increase | 63 [20] | 3·43 [1·82, 6·49] | **<0·001** | 37 [13] | 4·24 [1·99, 9·04] | **<0·001** | 26 [7] | 2·75 [0·70, 10·9] | 0·15 | 37 [13] | 3·61 [1·69, 5·94] | **<0·001** |
| Baseline SP-Ecog levels | 353 [52] | 1·15 [0·45, 2·97] | 0·80 | 215 [38] | 0·71 [0·23, 2·17] | 0·50 | 138 [14] | 10·4 [1·55, 69·8] | **0·02** | 215 [38] | 0·70 [0·46, 2·99] | 0·52 |
| Self-Ecog scores | | | **0·002** | | | 0·15 | | | 0·85 | | | 0·15 |
| Unchanged | 222 [27] | | | 143 [21] | | | 79 [6] | | | 143 [21] | | |
| Decrease | 64 [11] | 1·28 [0·59, 2·79] | 0·50 | 40 [7] | 1·19 [0·45, 3·14] | 0·70 | 24 [4] | 1·14 [0·23, 5·58] | 0·90 | 40 [7] | 0·99 [0·38, 2·61] | 0·99 |
| Increase | 67 [14] | 2·25 [1·13, 4·46] | **0·02** | 32 [10] | 2·93 [1·32, 6·48] | **0·008** | 35 [4] | 2·11 [0·45, 9·82] | 0·30 | 32 [10] | 2·18 [1·01, 4·70] | *0·047* |
| Baseline Self-Ecog levels | 353 [52] | 0·93 [0·36, 2·41] | 0·90 | 215 [38] | 1·05 [0·34, 3·21] | 0·90 | 138 [14] | 0·55 [0·06, 4·62] | 0·60 | 215 [38] | 1·12 [0·35, 3·60] | 0·85 |

**eTable 10.** Results from Models 2-3 replacing amyloid status by a combination of amyloid and tau status (AT status), including subgroup slopes and pairwise comparisons with Bonferroni-adjusted p-values using the ggeffects::hypothesis_test() function.

**eTable 8A**. Combined sample.

| | **SP-Ecog** | | | **Self-Ecog** | | | **Cum-Ecog** | | | **CAI-Ecog** | | |
|---|---|---|---|---|---|---|---|---|---|---|---|---|
| **Model 2 - Fixed Effect** | Est. | SE | P | Est. | SE | P | Est. | SE | P | Est. | SE | P |
| (Intercept) | 1·19 | 0·03 | **<0·001** | 1·22 | 0·03 | **<0·001** | 2·40 | 0·05 | **<0·001** | 0·02 | 0·04 | 0·57 |
| Time | 0·005 | 0·006 | 0·37 | 0·01 | 0·006 | *0·08* | 0·02 | 0·009 | *0·05* | 0·006 | 0·006 | 0·31 |
| AT status | | | 0·72 | | | 0·49 | | | 0·58 | | | 0·70 |
| Ab+T- | 0·02 | 0·03 | 0·41 | 0·03 | 0·03 | 0·39 | 0·03 | 0·04 | 0·41 | -0·007 | 0·03 | 0·84 |
| Ab+T+ | -0·003 | 0·03 | 0·92 | 0·04 | 0·04 | 0·33 | 0·05 | 0·06 | 0·40 | 0·03 | 0·04 | 0·45 |
| Time:AT status | | | **<0·001** | | | **<0·001** | | | **<0·001** | | | 0·23 |
| Ab+T- | 0·03 | 0·005 | **<0·001** | 0·02 | 0·006 | **0·002** | 0·04 | 0·008 | **<0·001** | -0·006 | 0·006 | 0·35 |
| Ab+T+ | 0·05 | 0·008 | **<0·001** | 0·03 | 0·008 | **<0·001** | 0·06 | 0·01 | **<0·001** | -0·01 | 0·008 | 0·11 |
| **Slopes** | Est. | 95% CI | $P_{adj}$ | Est. | 95% CI | $P_{adj}$ | Est. | 95% CI | $P_{adj}$ | Est. | 95% CI | $P_{adj}$ |
| Ab-T- | 0·004 | -0·007, 0·01 | 1·00 | 0·01 | -0·002, 0·02 | 0·30 | 0·02 | -0·002, 0·03 | 0·24 | 0·006 | -0·006, 0·02 | 0·91 |
| Ab+T- | 0·03 | 0·02, 0·04 | **<0·001** | 0·03 | 0·01, 0·04 | **<0·001** | 0·06 | 0·04, 0·08 | **<0·001** | 0·0009 | -0·01, 0·02 | 1·00 |
| Ab+T+ | 0·05 | 0·03, 0·07 | **<0·001** | 0·04 | 0·02, 0·06 | **<0·001** | 0·07 | 0·05, 0·10 | **<0·001** | -0·007 | -0·03, 0·01 | 1·00 |
| **Pairwise comparisons** | Est. | 95% CI | $P_{adj}$ | Slope | 95% CI | $P_{adj}$ | Slope | 95% CI | $P_{adj}$ | Slope | 95% CI | $P_{adj}$ |
| (Ab-T-)-(Ab+T-) | -0·03 | -0·04, -0·02 | **<0·001** | -0·02 | -0·03, -0·006 | **0·007** | -0·05 | -0·06, -0·03 | **<0·001** | 0·006 | -0·006, 0·02 | 1·00 |
| (Ab-T-)-(Ab+T+) | -0·05 | -0·06, -0·03 | **<0·001** | -0·03 | -0·05, -0·01 | **<0·001** | -0·06 | -0·08, -0·04 | **<0·001** | 0·01 | -0·003, 0·03 | 0·33 |
| (Ab+T-)-(Ab+T+) | -0·02 | -0·04, -0·003 | *0·06* | -0·01 | -0·03, 0·006 | 0·55 | -0·01 | -0·04, 0·01 | 0·88 | 0·008 | -0·01, 0·03 | 1·00 |
| **Model 3 - Fixed Effect** | Est. | SE | P | Est. | SE | P | Est. | SE | P | Est. | SE | P |
| (Intercept) | 1·18 | 0·03 | **<0·001** | 1·21 | 0·03 | **<0·001** | 2·38 | 0·05 | **<0·001** | 0·03 | 0·04 | 0·47 |
| Time | 0·005 | 0·005 | 0·31 | 0·01 | 0·006 | 0·10 | 0·02 | 0·008 | *0·05* | 0·005 | 0·006 | 0·37 |
| iMCI [iMCI] | 0·15 | 0·04 | **<0·001** | 0·08 | 0·05 | 0·10 | 0·23 | 0·06 | **<0·001** | -0·05 | 0·05 | 0·31 |
| AT status | | | 0·91 | | | 0·47 | | | 0·60 | | | 0·66 |
| Ab+T- | 0·01 | 0·03 | 0·62 | 0·02 | 0·03 | 0·48 | 0·02 | 0·05 | 0·59 | 0·001 | 0·04 | 0·97 |
| Ab+T+ | 0·003 | 0·04 | 0·94 | 0·05 | 0·05 | 0·28 | 0·06 | 0·07 | 0·34 | 0·05 | 0·05 | 0·38 |
| Time:iMCI [iMCI] | -0·0003 | 0·008 | 0·97 | 0·008 | 0·009 | 0·33 | 0·005 | 0·01 | 0·67 | 0·007 | 0·009 | 0·41 |
| Time:AT status | | | **0·008** | | | **0·001** | | | **0·001** | | | 0·29 |
| Time:Ab+T- | 0·01 | 0·006 | *0·02* | 0·006 | 0·006 | 0·32 | 0·02 | 0·008 | *0·02* | -0·006 | 0·006 | 0·37 |
| Time:Ab+T+ | 0·02 | 0·009 | *0·02* | 0·04 | 0·01 | **<0·001** | 0·04 | 0·01 | **0·002** | 0·01 | 0·01 | 0·26 |
| iMCI:AT status | | | 0·32 | | | 0·60 | | | 0·28 | | | 0·97 |
| Ab+T-.iMCI | -0·01 | 0·06 | 0·87 | -0·009 | 0·08 | 0·91 | -0·03 | 0·11 | 0·79 | -0·02 | 0·09 | 0·78 |

| | SP-Ecog | | | Self-Ecog | | | Cum-Ecog | | | CAI-Ecog | | |
|---|---|---|---|---|---|---|---|---|---|---|---|---|
| Ab+T+. iMCI | -0·11 | 0·07 | 0·13 | -0·09 | 0·09 | 0·32 | -0·19 | 0·12 | 0·11 | -0·007 | 0·10 | 0·95 |
| Time:iMCI:AT status | | | **<0·001** | | | **<0·001** | | | **<0·001** | | | **<0·001** |
| Ab+T-.iMCI | 0·07 | 0·01 | **<0·001** | 0·05 | 0·01 | **<0·001** | 0·13 | 0·02 | **<0·001** | -0·003 | 0·02 | 0·87 |
| Ab+T+.iMCI | 0·07 | 0·02 | **<0·001** | -0·02 | 0·02 | 0·21 | 0·06 | 0·02 | *0·02* | -0·08 | 0·02 | **<0·001** |
| **Slopes** | Est. | 95% CI | $P_{adj}$ | Est. | 95% CI | $P_{adj}$ | Est. | 95% CI | $P_{adj}$ | Est. | 95% CI | $P_{adj}$ |
| Ab-T-.Stable | 0·005 | -0·006, 0·01 | 1·00 | 0·009 | -0·002, 0·02 | 0·73 | 0·01 | -0·001, 0·03 | 0·43 | 0·006 | -0·006, 0·02 | 1·00 |
| Ab+T-.Stable | 0·02 | 0·004, 0·03 | *0·05* | 0·02 | 0·0006, 0·03 | 0·25 | 0·03 | 0·01, 0·06 | **0·004** | -0·0001 | -0·02, 0·02 | 1·00 |
| Ab+T+.Stable | 0·03 | 0·007, 0·05 | *0·05* | 0·05 | 0·02, 0·07 | **<0·001** | 0·05 | 0·03, 0·08 | **0·001** | 0·02 | -0·005, 0·04 | 0·77 |
| Ab-T-.iMCI | 0·004 | -0·01, 0·02 | 1·00 | 0·02 | -0·002, 0·04 | 0·43 | 0·02 | -0·006, 0·05 | 0·84 | 0·01 | -0·007, 0·03 | 1·00 |
| Ab+T-. iMCI | 0·09 | 0·07, 0·11 | **<0·001** | 0·08 | 0·05, 0·10 | **<0·001** | 0·17 | 0·13, 0·20 | **<0·001** | 0·005 | -0·02, 0·03 | 1·00 |
| Ab+T+. iMCI | 0·10 | 0·07, 0·12 | **<0·001** | 0·03 | 0·005, 0·06 | 0·13 | 0·11 | 0·08, 0·15 | **<0·001** | -0·05 | -0·08, -0·03 | **<0·001** |
| **Pairwise comparisons** | Est. | 95% CI | $P_{adj}$ | Slope | 95% CI | $P_{adj}$ | Slope | 95% CI | $P_{adj}$ | Slope | 95% CI | $P_{adj}$ |
| Ab-T-.Stable / Ab+T-.Stable | -0·01 | -0·02, -0·002 | 0·32 | -0·006 | -0·02, 0·006 | 1·00 | -0·02 | -0·04, -0·004 | 0·24 | 0·006 | -0·007, 0·02 | 1·00 |
| Ab-T-.Stable / Ab+T+.Stable | -0·02 | -0·04, -0·004 | 0·24 | -0·04 | -0·06, -0·02 | **0·003** | -0·04 | -0·06, -0·01 | **0·03** | -0·01 | -0·03, 0·008 | 1·00 |
| Ab-T-.Stable / Ab-T-.iMCI | 0·0003 | -0·01, 0·02 | 1·00 | -0·008 | -0·03, 0·009 | 1·00 | -0·005 | -0·03, 0·02 | 1·00 | -0·007 | -0·02, 0·01 | 1·00 |
| Ab-T-.Stable / Ab+T-.iMCI | -0·08 | -0·10, -0·06 | **<0·001** | -0·07 | -0·09, -0·05 | **<0·001** | -0·15 | -0·18, -0·12 | **<0·001** | 0·001 | -0·02, 0·02 | 1·00 |
| Ab-T-.Stable / Ab+T+.iMCI | -0·09 | -0·12, -0·07 | **<0·001** | -0·02 | -0·05, 0·003 | 1·00 | -0·10 | -0·13, -0·07 | **<0·001** | 0·06 | 0·03, 0·09 | **<0·001** |
| Ab+T-.Stable / Ab+T+.Stable | -0·009 | -0·03, 0·01 | 1·00 | -0·03 | -0·05, -0·009 | *0·08* | -0·02 | -0·05, 0·009 | 1·00 | -0·02 | -0·04, 0·004 | 1·00 |
| Ab+T-.Stable / Ab-T-.iMCI | 0·01 | -0·004, 0·03 | 1·00 | -0·002 | -0·02, 0·02 | 1·00 | 0·02 | -0·01, 0·04 | 1·00 | -0·01 | -0·03, 0·007 | 1·00 |
| Ab+T-.Stable / Ab+T-.iMCI | -0·07 | -0·09, -0·05 | **<0·001** | -0·06 | -0·09, -0·04 | **<0·001** | -0·13 | -0·17, -0·10 | **<0·001** | -0·005 | -0·03, 0·02 | 1·00 |
| Ab+T-.Stable / Ab+T+.iMCI | -0·08 | -0·11, -0·06 | **<0·001** | -0·02 | -0·04, 0·01 | 1·00 | -0·08 | -0·12, -0·04 | **<0·001** | 0·05 | 0·03, 0·08 | **0·002** |
| Ab+T+.Stable / Ab-T-.iMCI | 0·02 | 0·0000, 0·04 | 0·74 | 0·03 | 0·004, 0·05 | 0·35 | 0·03 | 0·002, 0·07 | 0·58 | 0·004 | -0·02, 0·03 | 1·00 |
| Ab+T+.Stable / Ab+T-.iMCI | -0·06 | -0·09, -0·03 | **<0·001** | -0·03 | -0·06, -0·003 | 0·41 | -0·11 | -0·15, -0·07 | **<0·001** | 0·01 | -0·02, 0·04 | 1·00 |
| Ab+T+.Stable / Ab+T+.iMCI | -0·07 | -0·10, -0·04 | **<0·001** | 0·01 | -0·02, 0·04 | 1·00 | -0·06 | -0·10, -0·02 | *0·05* | 0·07 | 0·04, 0·10 | **<0·001** |
| Ab-T-.iMCI / Ab+T-.iMCI | -0·08 | -0·11, -0·06 | **<0·001** | -0·06 | -0·09, -0·03 | **<0·001** | -0·15 | -0·18, -0·11 | **<0·001** | 0·008 | -0·02, 0·04 | 1·00 |
| Ab-T-.iMCI / Ab+T+.iMCI | -0·09 | -0·12, -0·07 | **<0·001** | -0·01 | -0·04, 0·02 | 1·00 | -0·09 | -0·13, -0·06 | **<0·001** | 0·07 | 0·04, 0·10 | **<0·001** |
| Ab+T-.iMCI / Ab+T+.iMCI | -0·01 | -0·04, 0·02 | 1·00 | 0·05 | 0·01, 0·08 | *0·09* | 0·05 | 0·008, 0·10 | 0·32 | 0·06 | 0·02, 0·09 | **0·010** |

To reduce the number of lines, those referring to covariates are not presented above. Adjusted for age, sex, years of education, amyloid and tau modalities (PET, CSF, plasma, Mixed), cohort (only for combined analyses), setting (only for DELCODE, except when restricted to SCD patients), and their interaction with time.

**eTable 8B**. DELCODE cohort.

| | **SP-Ecog** | | | **Self-Ecog** | | | **Cum-Ecog** | | | **CAI-Ecog** | | |
|---|---|---|---|---|---|---|---|---|---|---|---|---|
| **Model 2 - Fixed Effect** | Est. | SE | P | Est. | SE | P | Est. | SE | P | Est. | SE | P |
| (Intercept) | 1·21 | 0·03 | **<0·001** | 1·24 | 0·03 | **<0·001** | 2·44 | 0·04 | **<0·001** | 0·02 | 0·04 | 0·53 |
| Time | 0·007 | 0·005 | 0·18 | 0·01 | 0·006 | *0·09* | 0·02 | 0·009 | *0·03* | 0·007 | 0·006 | 0·29 |
| AT status | | | 0·67 | | | 0·83 | | | 0·80 | | | 0·93 |
| Ab+T- | -0·006 | 0·03 | 0·92 | 0·01 | 0·04 | 0·71 | -0·003 | 0·05 | 0·96 | 0·004 | 0·04 | 0·93 |
| Ab+T+ | -0·04 | 0·05 | 0·37 | -0·02 | 0·06 | 0·69 | -0·05 | 0·08 | 0·51 | 0·03 | 0·07 | 0·71 |
| Time:AT status | | | **<0·001** | | | **0·008** | | | **<0·001** | | | **0·008** |
| Ab+T- | 0·03 | 0·006 | **<0·001** | 0·02 | 0·007 | **0·008** | 0·05 | 0·01 | **<0·001** | -0·01 | 0·007 | *0·05* |
| Ab+T+ | 0·05 | 0·01 | **<0·001** | 0·03 | 0·01 | *0·03* | 0·07 | 0·02 | **<0·001** | -0·04 | 0·01 | **0·006** |
| **Slopes** | Est. | 95% CI | $P_{adj}$ | Est. | 95% CI | $P_{adj}$ | Est. | 95% CI | $P_{adj}$ | Est. | 95% CI | $P_{adj}$ |
| Ab-T- | 0·007 | -0·004, 0·02 | 0·64 | 0·009 | -0·002, 0·02 | 0·35 | 0·02 | 0·0004, 0·04 | 0·14 | 0·007 | -0·006, 0·02 | 0·93 |
| Ab+T- | 0·04 | 0·03, 0·05 | **<0·001** | 0·03 | 0·01, 0·04 | **<0·001** | 0·07 | 0·05, 0·09 | **<0·001** | -0·008 | -0·02, 0·007 | 0·95 |
| Ab+T+ | 0·06 | 0·04, 0·08 | **<0·001** | 0·03 | 0·01, 0·06 | **0·01** | 0·09 | 0·05, 0·13 | **<0·001** | -0·03 | -0·06,-0·004 | *0·07* |
| **Pairwise comparisons** | Est. | 95% CI | $P_{adj}$ | Slope | 95% CI | $P_{adj}$ | Slope | 95% CI | $P_{adj}$ | Slope | 95% CI | $P_{adj}$ |
| (Ab-T-)-(Ab+T-) | -0·03 | -0·04, -0·02 | **<0·001** | -0·02 | -0·03, -0·005 | **0·02** | -0·05 | -0·07, -0·03 | **<0·001** | 0·01 | 0·0001, 0·03 | 0·15 |
| (Ab-T-)-(Ab+T+) | -0·05 | -0·07, -0·03 | **<0·001** | -0·03 | -0·05, -0·003 | *0·09* | -0·07 | -0·11, -0·03 | **<0·001** | 0·04 | 0·01, 0·06 | **0·02** |
| (Ab+T-)-(Ab+T+) | -0·02 | -0·04, 0·002 | 0·23 | -0·008 | -0·03, 0·02 | 1·00 | -0·02 | -0·06, 0·02 | 1·00 | 0·02 | -0·005, 0·05 | 0·33 |
| **Model 3 - Fixed Effect** | Est. | SE | P | Est. | SE | P | Est. | SE | P | Est. | SE | P |
| (Intercept) | 1·20 | 0·03 | **<0·001** | 1·24 | 0·03 | **<0·001** | 2·42 | 0·04 | **<0·001** | 0·02 | 0·04 | 0·56 |
| Time | 0·008 | 0·005 | 0·11 | 0·008 | 0·006 | 0·15 | 0·02 | 0·009 | *0·02* | 0·005 | 0·007 | 0·44 |
| iMCI [iMCI] | 0·09 | 0·04 | *0·04* | 0·05 | 0·05 | 0·28 | 0·16 | 0·07 | *0·02* | -0·01 | 0·06 | 0·86 |
| AT status | | | 0·48 | | | 0·97 | | | 0·89 | | | 0·55 |
| Ab+T- | -0·02 | 0·03 | 0·48 | 0·01 | 0·04 | 0·80 | -0·02 | 0·06 | 0·72 | 0·03 | 0·05 | 0·55 |
| Ab+T+ | -0·07 | 0·06 | 0·27 | 0·004 | 0·08 | 0·95 | -0·04 | 0·10 | 0·69 | 0·09 | 0·09 | 0·31 |
| Time:iMCI [iMCI] | 0·003 | 0·008 | 0·72 | 0·02 | 0·009 | *0·07* | 0·02 | 0·01 | 0·24 | 0·009 | 0·01 | 0·38 |
| Time:AT status | | | *0·015* | | | 0·17 | | | *0·05* | | | *0·32* |
| Time:Ab+T- | 0·01 | 0·006 | *0·02* | 0·007 | 0·007 | 0·35 | 0·02 | 0·01 | *0·04* | -0·01 | 0·008 | 0·19 |
| Time:Ab+T+ | 0·03 | 0·01 | *0·04* | 0·03 | 0·01 | *0·08* | 0·04 | 0·02 | *0·09* | -0·02 | 0·02 | 0·35 |
| iMCI:AT status | | | 0·68 | | | 0·72 | | | 0·70 | | | 0·46 |
| Ab+T-.iMCI | 0·06 | 0·07 | 0·39 | 0·003 | 0·09 | 0·98 | 0·03 | 0·12 | 0·81 | -0·10 | 0·11 | 0·33 |
| Ab+T+. iMCI | 0·007 | 0·10 | 0·94 | -0·09 | 0·12 | 0·44 | -0·12 | 0·16 | 0·46 | -0·14 | 0·14 | 0·33 |
| Time:iMCI:AT status | | | **<0·001** | | | *0·03* | | | **<0·001** | | | *0·10* |
| Ab+T-.iMCI | 0·08 | 0·01 | **<0·001** | 0·04 | 0·02 | **0·012** | 0·13 | 0·02 | **<0·001** | -0·02 | 0·02 | 0·26 |

| | SP-Ecog | | | Self-Ecog | | | Cum-Ecog | | | CAI-Ecog | | |
|---|---|---|---|---|---|---|---|---|---|---|---|---|
| Ab+T+.iMCI | 0·06 | 0·02 | **0·004** | -0·008 | 0·02 | 0·73 | 0·07 | 0·03 | *0·05* | -0·06 | 0·03 | *0·04* |
| **Slopes** | Est. | 95% CI | $P_{adj}$ | Est. | 95% CI | $P_{adj}$ | Est. | 95% CI | $P_{adj}$ | Est. | 95% CI | $P_{adj}$ |
| Ab-T-.Stable | 0·008 | -0·002, 0·02 | 0·79 | 0·008 | -0·004, 0·02 | 1·00 | 0·02 | 0·002, 0·04 | 0·19 | 0·005 | -0·008, 0·02 | 1·00 |
| Ab+T-.Stable | 0·02 | 0·01, 0·04 | **0·004** | 0·01 | -0·0004, 0·03 | 0·34 | 0·04 | 0·02, 0·06 | **0·002** | -0·006 | -0·02, 0·01 | 1·00 |
| Ab+T+.Stable | 0·03 | 0·009, 0·06 | **0·04** | 0·03 | 0·005, 0·06 | 0·12 | 0·06 | 0·01, 0·10 | *0·06* | -0·01 | -0·04, 0·02 | 1·00 |
| Ab-T-.iMCI | 0·01 | -0·006, 0·03 | 1·00 | 0·02 | 0·005, 0·04 | *0·08* | 0·04 | 0·006, 0·06 | 0·11 | 0·01 | -0·008, 0·04 | 1·00 |
| Ab+T-. iMCI | 0·10 | 0·08, 0·12 | **<0·001** | 0·07 | 0·05, 0·10 | **<0·001** | 0·19 | 0·15, 0·22 | **<0·001** | -0·02 | -0·05, 0·01 | 1·00 |
| Ab+T+. iMCI | 0·10 | 0·07, 0·13 | **<0·001** | 0·04 | 0·008, 0·08 | 0·10 | 0·14 | 0·09, 0·19 | **<0·001** | -0·06 | -0·10, -0·02 | **0·02** |
| **Pairwise comparisons** | Est. | 95% CI | $P_{adj}$ | Slope | 95% CI | $P_{adj}$ | Slope | 95% CI | $P_{adj}$ | Slope | 95% CI | $P_{adj}$ |
| Ab-T-.Stable / Ab+T-.Stable | -0·01 | -0·03, -0·002 | 0·28 | -0·007 | -0·02, 0·008 | 1·00 | -0·02 | -0·04,-0·0006 | 0·66 | 0·01 | -0·005, 0·03 | 1·00 |
| Ab-T-.Stable / Ab+T+.Stable | -0·03 | -0·05, -0·002 | 0·56 | -0·03 | -0·05, 0·003 | 1·00 | -0·04 | -0·08, 0·005 | 1·00 | 0·02 | -0·02, 0·05 | 1·00 |
| Ab-T-.Stable / Ab-T-.iMCI | -0·003 | -0·02, 0·01 | 1·00 | -0·02 | -0·04, 0·002 | 1·00 | -0·02 | -0·04, 0·01 | 1·00 | -0·009 | -0·03, 0·01 | 1·00 |
| Ab-T-.Stable / Ab+T-.iMCI | -0·09 | -0·12, -0·07 | **<0·001** | -0·06 | -0·09, -0·04 | **<0·001** | -0·17 | -0·20, -0·13 | **<0·001** | 0·02 | -0·005, 0·05 | 1·00 |
| Ab-T-.Stable / Ab+T+.iMCI | -0·09 | -0·12, -0·06 | **<0·001** | -0·03 | -0·07, -0·0003 | 0·72 | -0·12 | -0·17, -0·07 | **<0·001** | 0·06 | 0·02, 0·10 | **0·02** |
| Ab+T-.Stable / Ab+T+.Stable | -0·01 | -0·04, 0·01 | 1·00 | -0·02 | -0·05, 0·01 | 1·00 | -0·02 | -0·06, 0·03 | 1·00 | 0·005 | -0·03, 0·04 | 1·00 |
| Ab+T-.Stable / Ab-T-.iMCI | 0·01 | -0·006, 0·03 | 1·00 | -0·01 | -0·03, 0·01 | 1·00 | 0·006 | -0·03, 0·04 | 1·00 | -0·02 | -0·04, 0·004 | 1·00 |
| Ab+T-.Stable / Ab+T-.iMCI | -0·08 | -0·10, -0·06 | **<0·001** | -0·06 | -0·08, -0·03 | **<0·001** | -0·15 | -0·19, -0·11 | **<0·001** | 0·01 | -0·02, 0·04 | 1·00 |
| Ab+T-.Stable / Ab+T+.iMCI | -0·07 | -0·10, -0·04 | **<0·001** | -0·03 | -0·06, 0·008 | 1·00 | -0·10 | -0·15, -0·05 | **0·002** | 0·05 | 0·01, 0·09 | 0·17 |
| Ab+T+.Stable / Ab-T-.iMCI | 0·02 | -0·004, 0·05 | 1·00 | 0·008 | -0·02, 0·04 | 1·00 | 0·02 | -0·03, 0·07 | 1·00 | -0·03 | -0·06, 0·01 | 1·00 |
| Ab+T+.Stable / Ab+T-.iMCI | -0·07 | -0·10, -0·04 | **<0·001** | -0·04 | -0·07, -0·004 | 0·41 | -0·13 | -0·18, -0·08 | **<0·001** | 0·006 | -0·04, 0·05 | 1·00 |
| Ab+T+.Stable / Ab+T+.iMCI | -0·06 | -0·10, -0·03 | **0·01** | -0·009 | -0·05, 0·03 | 1·00 | -0·09 | -0·15, -0·02 | 0·11 | 0·05 | -0·002, 0·10 | 0·94 |
| Ab-T-.iMCI / Ab+T-.iMCI | -0·09 | -0·12, -0·07 | **<0·001** | -0·05 | -0·08, -0·02 | **0·01** | -0·15 | -0·19, -0·11 | **<0·001** | 0·03 | -0·001, 0·06 | 0·87 |
| Ab-T-.iMCI / Ab+T+.iMCI | -0·09 | -0·12, -0·05 | **<0·001** | -0·02 | -0·05, 0·02 | 1·00 | -0·11 | -0·16, -0·05 | **0·002** | 0·07 | 0·03, 0·11 | **0·01** |
| Ab+T-.iMCI / Ab+T+.iMCI | 0·006 | -0·03, 0·04 | 1·00 | 0·03 | -0·009, 0·07 | 1·00 | 0·04 | -0·02, 0·10 | 1·00 | 0·04 | -0·005, 0·09 | 1·00 |

**eTable 8C**. DELCODE cohort restricted to SCD patients.

| | **SP-Ecog** | | | **Self-Ecog** | | | **Cum-Ecog** | | | **CAI-Ecog** | | |
|---|---|---|---|---|---|---|---|---|---|---|---|---|
| **Model 2 - Fixed Effect** | Est. | SE | P | Est. | SE | P | Est. | SE | P | Est. | SE | P |
| (Intercept) | 1·45 | 0·04 | **<0·001** | 1·55 | 0·04 | **<0·001** | 2·98 | 0·06 | **<0·001** | 0·09 | 0·05 | 0·12 |
| Time | 0·02 | 0·008 | *0·04* | 0·007 | 0·009 | 0·45 | 0·03 | 0·01 | *0·04* | 0·002 | 0·009 | 0·86 |
| AT status | | | 0·75 | | | 0·95 | | | 0·82 | | | 0·82 |
| Ab+T- | -0·02 | 0·05 | 0·75 | -0·02 | 0·06 | 0·78 | -0·05 | 0·08 | 0·53 | -0·02 | 0·08 | 0·78 |
| Ab+T+ | -0·05 | 0·07 | 0·46 | 0·006 | 0·08 | 0·94 | -0·02 | 0·11 | 0·83 | 0·06 | 0·10 | 0·53 |
| Time:AT status | | | **<0·001** | | | *0·07* | | | **<0·001** | | | **<0·001** |
| Ab+T- | 0·05 | 0·010 | **<0·001** | 0·02 | 0·01 | *0·05* | 0·07 | 0·02 | **<0·001** | -0·02 | 0·01 | *0·08* |
| Ab+T+ | 0·07 | 0·02 | **<0·001** | 0·03 | 0·02 | 0·10 | 0·09 | 0·03 | **0·001** | -0·06 | 0·02 | **0·002** |
| **Slopes** | Est. | 95% CI | $P_{adj}$ | Est. | 95% CI | $P_{adj}$ | Est. | 95% CI | $P_{adj}$ | Est. | 95% CI | $P_{adj}$ |
| Ab-T- | 0·02 | 0·002, 0·03 | *0·08* | 0·009 | -0·008, 0·03 | 0·86 | 0·03 | 0·005, 0·06 | *0·06* | 0·003 | -0·02, 0·02 | 1·00 |
| Ab+T- | 0·06 | 0·04, 0·08 | **<0·001** | 0·03 | 0·01, 0·05 | **0·01** | 0·11 | 0·07, 0·14 | **<0·001** | -0·02 | -0·04, 0·004 | 0·33 |
| Ab+T+ | 0·09 | 0·06, 0·12 | **<0·001** | 0·04 | 0·005, 0·07 | *0·07* | 0·12 | 0·07, 0·17 | **<0·001** | -0·06 | -0·09, -0·02 | **0·005** |
| **Pairwise comparisons** | Est. | 95% CI | $P_{adj}$ | Slope | 95% CI | $P_{adj}$ | Slope | 95% CI | $P_{adj}$ | Slope | 95% CI | $P_{adj}$ |
| (Ab-T-)-(Ab+T-) | -0·05 | -0·07, -0·03 | **<0·001** | -0·02 | -0·05, -0·0006 | 0·13 | -0·07 | -0·11, -0·04 | **<0·001** | 0·02 | -0·002, 0·05 | 0·23 |
| (Ab-T-)-(Ab+T+) | -0·07 | -0·10, -0·04 | **<0·001** | -0·03 | -0·06, 0·005 | 0·31 | -0·09 | -0·14, -0·03 | **0·004** | 0·06 | 0·02, 0·10 | **0·005** |
| (Ab+T-)-(Ab+T+) | -0·02 | -0·06, 0·007 | 0·38 | -0·004 | -0·04, 0·03 | 1·00 | -0·01 | -0·07, 0·04 | 1·00 | 0·04 | -0·002, 0·08 | 0·19 |
| **Model 3 - Fixed Effect** | Est. | SE | P | Est. | SE | P | Est. | SE | P | Est. | SE | P |
| (Intercept) | 1·42 | 0·04 | **<0·001** | 1·54 | 0·05 | **<0·001** | 2·94 | 0·06 | **<0·001** | 0·09 | 0·06 | 0·13 |
| Time | 0·01 | 0·008 | *0·08* | 0·001 | 0·009 | 0·90 | 0·02 | 0·01 | 0·10 | -0·0007 | 0·01 | 0·95 |
| iMCI [iMCI] | 0·10 | 0·06 | 0·12 | 0·05 | 0·08 | 0·53 | 0·17 | 0·10 | *0·09* | -0·02 | 0·09 | 0·85 |
| AT status | | | 0·70 | | | 0·91 | | | 0·94 | | | 0·55 |
| Ab+T- | -0·03 | 0·06 | 0·62 | 0·01 | 0·07 | 0·87 | -0·03 | 0·09 | 0·78 | 0·03 | 0·09 | 0·73 |
| Ab+T+ | -0·07 | 0·09 | 0·44 | 0·05 | 0·11 | 0·66 | 0·02 | 0·15 | 0·89 | 0·15 | 0·14 | 0·28 |
| Time:iMCI [iMCI] | 0·007 | 0·01 | 0·60 | 0·03 | 0·01 | 0·08 | 0·03 | 0·02 | 0·19 | 0·01 | 0·02 | 0·46 |
| Time:AT status | | | *0·014* | | | 0·30 | | | 0·11 | | | 0·21 |
| Time:Ab+T- | 0·02 | 0·01 | *0·03* | 0·010 | 0·01 | 0·46 | 0·03 | 0·02 | *0·09* | -0·02 | 0·01 | 0·23 |
| Time:Ab+T+ | 0·05 | 0·02 | *0·02* | 0·03 | 0·02 | 0·14 | 0·05 | 0·03 | 0·12 | -0·04 | 0·03 | 0·14 |
| iMCI:AT status | | | 0·97 | | | 0·67 | | | 0·66 | | | 0·52 |
| Ab+T-.iMCI | 0·02 | 0·11 | 0·83 | -0·10 | 0·13 | 0·46 | -0·11 | 0·17 | 0·52 | -0·16 | 0·17 | 0·33 |
| Ab+T+. iMCI | -0·009 | 0·14 | 0·95 | -0·11 | 0·17 | 0·51 | -0·17 | 0·22 | 0·43 | -0·17 | 0·21 | 0·42 |
| Time:iMCI:AT status | | | **0·002** | | | 0·19 | | | **0·005** | | | 0·45 |
| Ab+T-.iMCI | 0·08 | 0·02 | **<0·001** | 0·04 | 0·02 | 0·15 | 0·13 | 0·04 | **0·001** | -0·02 | 0·03 | 0·49 |

| | SP-Ecog | | | Self-Ecog | | | Cum-Ecog | | | CAI-Ecog | | |
|---|---|---|---|---|---|---|---|---|---|---|---|---|
| Ab+T+.iMCI | 0·05 | 0·03 | 0·11 | -0·02 | 0·03 | 0·48 | 0·05 | 0·05 | 0·32 | -0·05 | 0·04 | 0·23 |
| **Slopes** | Est. | 95% CI | $P_{adj}$ | Est. | 95% CI | $P_{adj}$ | Est. | 95% CI | $P_{adj}$ | Est. | 95% CI | $P_{adj}$ |
| Ab-T-.Stable | 0·01 | -0·0003, 0·03 | 0·33 | 0·003 | -0·01, 0·02 | 1·00 | 0·03 | -0·001, 0·05 | 0·38 | 0·0004 | -0·02, 0·02 | 1·00 |
| Ab+T-.Stable | 0·04 | 0·02, 0·06 | **0·001** | 0·01 | -0·01, 0·04 | 1·00 | 0·06 | 0·02, 0·10 | **0·01** | -0·02 | -0·04, 0·010 | 1·00 |
| Ab+T+.Stable | 0·06 | 0·02, 0·10 | **0·01** | 0·04 | -0·006, 0·08 | 0·55 | 0·08 | 0·01, 0·15 | 0·11 | -0·04 | -0·09, 0·01 | 0·77 |
| Ab-T-.iMCI | 0·02 | -0·003, 0·05 | 0·49 | 0·03 | 0·001, 0·06 | 0·25 | 0·06 | 0·01, 0·10 | *0·07* | 0·01 | -0·02, 0·04 | 1·00 |
| Ab+T-. iMCI | 0·12 | 0·09, 0·15 | **<0·001** | 0·08 | 0·04, 0·11 | **<0·001** | 0·22 | 0·16, 0·27 | **<0·001** | -0·02 | -0·06, 0·02 | 1·00 |
| Ab+T+. iMCI | 0·11 | 0·08, 0·15 | **<0·001** | 0·04 | -0·005, 0·08 | 0·51 | 0·16 | 0·09, 0·22 | **<0·001** | -0·07 | -0·12, -0·02 | **0·02** |
| **Pairwise comparisons** | Est. | 95% CI | $P_{adj}$ | Slope | 95% CI | $P_{adj}$ | Slope | 95% CI | $P_{adj}$ | Slope | 95% CI | $P_{adj}$ |
| Ab-T-.Stable / Ab+T-.Stable | -0·02 | -0·05, -0·002 | 0·44 | -0·010 | -0·04, 0·02 | 1·00 | -0·03 | -0·07, 0·005 | 1·00 | 0·02 | -0·01, 0·05 | 1·00 |
| Ab-T-.Stable / Ab+T+.Stable | -0·05 | -0·08, -0·007 | 0·31 | -0·03 | -0·07, 0·01 | 1·00 | -0·05 | -0·12, 0·01 | 1·00 | 0·04 | -0·01, 0·09 | 1·00 |
| Ab-T-.Stable / Ab-T-.iMCI | -0·007 | -0·03, 0·02 | 1·00 | -0·03 | -0·06, 0·003 | 1·00 | -0·03 | -0·07, 0·01 | 1·00 | -0·01 | -0·05, 0·02 | 1·00 |
| Ab-T-.Stable / Ab+T-.iMCI | -0·11 | -0·14, -0·08 | **<0·001** | -0·07 | -0·11, -0·04 | **0·001** | -0·19 | -0·25, -0·13 | **<0·001** | 0·02 | -0·02, 0·07 | 1·00 |
| Ab-T-.Stable / Ab+T+.iMCI | -0·10 | -0·14, -0·06 | **<0·001** | -0·03 | -0·08, 0·01 | 1·00 | -0·13 | -0·20, -0·07 | **0·002** | 0·07 | 0·02, 0·12 | *0·07* |
| Ab+T-.Stable / Ab+T+.Stable | -0·02 | -0·06, 0·02 | 1·00 | -0·02 | -0·07, 0·02 | 1·00 | -0·02 | -0·09, 0·05 | 1·00 | 0·02 | -0·03, 0·08 | 1·00 |
| Ab+T-.Stable / Ab-T-.iMCI | 0·02 | -0·01, 0·05 | 1·00 | -0·02 | -0·05, 0·02 | 1·00 | 0·004 | -0·05, 0·06 | 1·00 | -0·03 | -0·07, 0·009 | 1·00 |
| Ab+T-.Stable / Ab+T-.iMCI | -0·08 | -0·12, -0·05 | **<0·001** | -0·06 | -0·10, -0·02 | **0·03** | -0·16 | -0·22, -0·09 | **<0·001** | 0·007 | -0·04, 0·05 | 1·00 |
| Ab+T-.Stable / Ab+T+.iMCI | -0·08 | -0·12, -0·04 | **0·003** | -0·02 | -0·07, 0·02 | 1·00 | -0·10 | -0·17, -0·03 | *0·09* | 0·05 | 0·002, 0·11 | 0·63 |
| Ab+T+.Stable / Ab-T-.iMCI | 0·04 | -0·004, 0·08 | 1·00 | 0·006 | -0·04, 0·05 | 1·00 | 0·02 | -0·05, 0·10 | 1·00 | -0·05 | -0·11, 0·005 | 1·00 |
| Ab+T+.Stable / Ab+T-.iMCI | -0·06 | -0·11, -0·01 | 0·16 | -0·04 | -0·09, 0·01 | 1·00 | -0·14 | -0·22, -0·05 | **0·02** | -0·01 | -0·08, 0·05 | 1·00 |
| Ab+T+.Stable / Ab+T+.iMCI | -0·05 | -0·10, -0·002 | 0·61 | -0·003 | -0·06, 0·06 | 1·00 | -0·08 | -0·17, 0·010 | 1·00 | 0·03 | -0·03, 0·10 | 1·00 |
| Ab-T-.iMCI / Ab+T-.iMCI | -0·10 | -0·14, -0·06 | **<0·001** | -0·05 | -0·09, -0·004 | 0·46 | -0·16 | -0·23, -0·10 | **<0·001** | 0·04 | -0·01, 0·09 | 1·00 |
| Ab-T-.iMCI / Ab+T+.iMCI | -0·09 | -0·13, -0·05 | **<0·001** | -0·009 | -0·06, 0·04 | 1·00 | -0·10 | -0·18, -0·03 | *0·10* | 0·09 | 0·03, 0·14 | **0·04** |
| Ab+T-.iMCI / Ab+T+.iMCI | 0·007 | -0·04, 0·05 | 1·00 | 0·04 | -0·02, 0·09 | 1·00 | 0·06 | -0·03, 0·14 | 1·00 | 0·05 | -0·01, 0·11 | 1·00 |

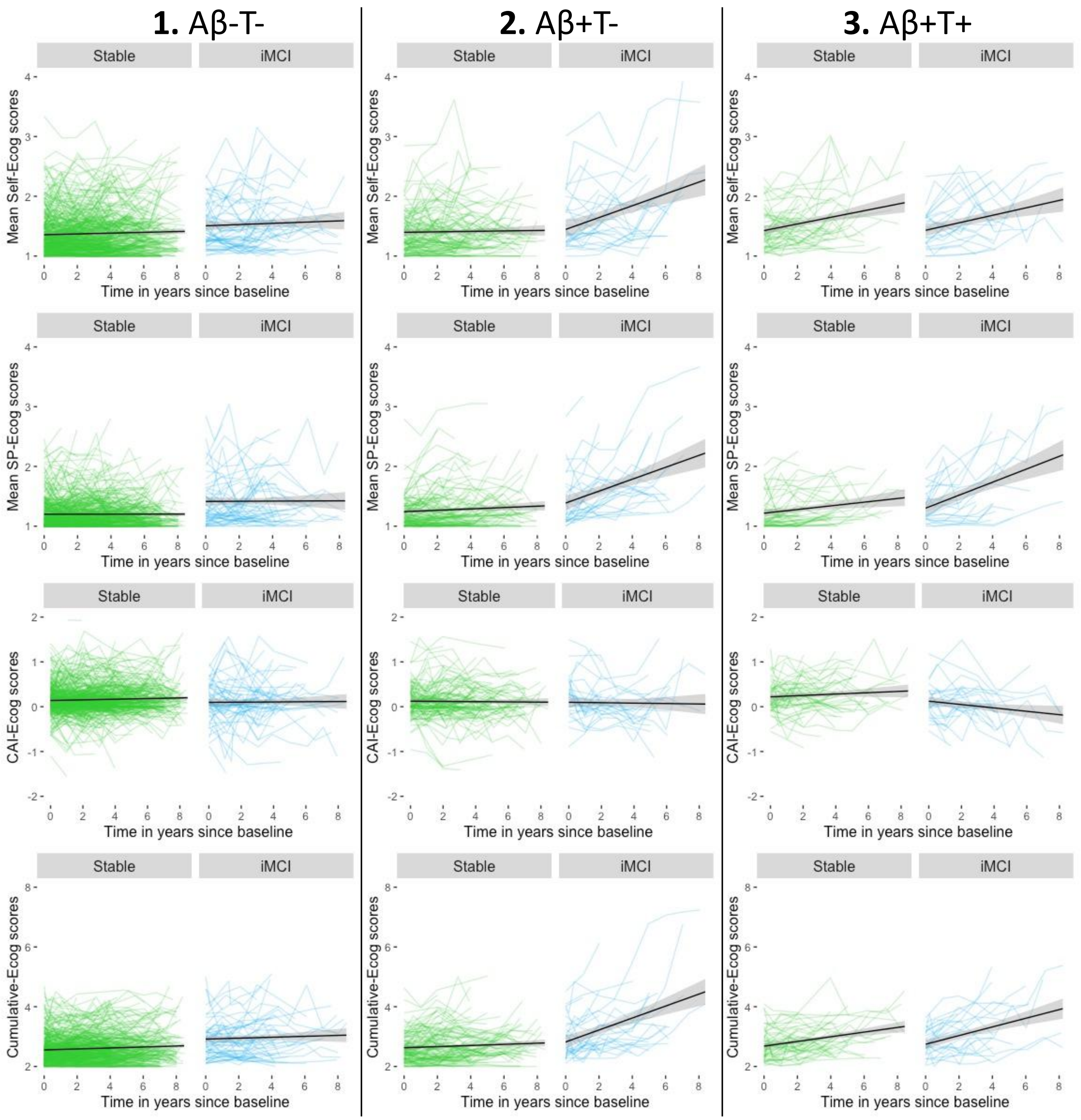

**eFigure 2.** Spaghetti plot of individual trajectories: SCD reports over time in years since baseline according to clinical progression and AD pathology (amyloid and tau status).

Abbreviations: Aβ-T-, participants amyloid- and tau-negative at baseline; Aβ+T-, participants amyloid-positive but tau-negative at baseline; Aβ+T+, participants amyloid- and tau-positive at baseline; CAI, Cognitive awareness index; iMCI, participants that progressed to incident mild cognitive impairment during the follow-up period; SP, study partner; Stable, participants cognitively stable during the follow-up period.

#### References:

1. Jessen F, Spottke A, Boecker H, et al. Design and first baseline data of the DZNE multicenter observational study on predementia Alzheimer's disease (DELCODE). *Alzheimers Res Ther*. 02 2018;10(1):15. doi:10.1186/s13195-017-0314-2
2. Jessen F, Amariglio RE, van Boxtel M, et al. A conceptual framework for research on subjective cognitive decline in preclinical Alzheimer's disease. *Alzheimer's & Dementia: The Journal of the Alzheimer's Association*. 2014/11// 2014;10(6):844-852. doi:10.1016/j.jalz.2014.01.001
3. Molinuevo JL, Rabin LA, Amariglio R, et al. Implementation of subjective cognitive decline criteria in research studies. *Alzheimer's & Dementia: The Journal of the Alzheimer's Association*. 2017/03// 2017;13(3):296-311. doi:10.1016/j.jalz.2016.09.012
4. Stark M, Wolfsgruber S, Kleineidam L, et al. Relevance of Minor Neuropsychological Deficits in Patients With Subjective Cognitive Decline. *Neurology*. Nov 21 2023;101(21):e2185-e2196. doi:10.1212/WNL.0000000000207844
5. Winblad B, Palmer K, Kivipelto M, et al. Mild cognitive impairment--beyond controversies, towards a consensus: report of the International Working Group on Mild Cognitive Impairment. *J Intern Med*. Sep 2004;256(3):240-6. doi:10.1111/j.1365-2796.2004.01380.x
6. Albert MS, DeKosky ST, Dickson D, et al. The diagnosis of mild cognitive impairment due to Alzheimer's disease: recommendations from the National Institute on Aging-Alzheimer's Association workgroups on diagnostic guidelines for Alzheimer's disease. *Alzheimers Dement*. May 2011;7(3):270-9. doi:10.1016/j.jalz.2011.03.008
7. Bertens D, Tijms BM, Scheltens P, Teunissen CE, Visser PJ. Unbiased estimates of cerebrospinal fluid β-amyloid 1-42 cutoffs in a large memory clinic population. *Alzheimers Res Ther*. Feb 14 2017;9(1):8. doi:10.1186/s13195-016-0233-7
8. Jessen F, Wolfsgruber S, Kleineindam L, et al. Subjective cognitive decline and stage 2 of Alzheimer disease in patients from memory centers. *Alzheimers Dement*. Feb 2023;19(2):487-497. doi:10.1002/alz.12674
9. Vogelgsang J, Hansen N, Stark M, et al. Plasma amyloid beta X-42/X-40 ratio and cognitive decline in suspected early and preclinical Alzheimer's disease. *Alzheimers Dement*. Aug 2024;20(8):5132-5142. doi:10.1002/alz.13909
10. Mengel D, Soter E, Ott JM, et al. Blood biomarkers confirm subjective cognitive decline (SCD) as a distinct molecular and clinical stage within the NIA-AA framework of Alzheimer's disease. *medRxiv*. 2024:2024.07.10.24310205. doi:10.1101/2024.07.10.24310205
11. Weiner MW, Veitch DP, Aisen PS, et al. 2014 Update of the Alzheimer's Disease Neuroimaging Initiative: A review of papers published since its inception. *Alzheimer's & dementia : the journal of the Alzheimer's Association*. 2015/06// 2015;11(6):e1-120. doi:10.1016/j.jalz.2014.11.001
12. Folstein MF, Folstein SE, McHugh PR. "Mini-mental state": A practical method for grading the cognitive state of patients for the clinician. *Journal of Psychiatric Research*. 1975/11/01/ 1975;12(3):189-198. doi:10.1016/0022-3956(75)90026-6
13. Morris JC. The Clinical Dementia Rating (CDR): current version and scoring rules. *Neurology*. Nov 1993;43(11):2412-4. doi:10.1212/wnl.43.11.2412-a
14. Saykin AJ, Wishart HA, Rabin LA, et al. Older adults with cognitive complaints show brain atrophy similar to that of amnestic MCI. *Neurology*. 2006/09/12/ 2006;67(5):834-842. doi:10.1212/01.wnl.0000234032.77541.a2
15. Landau SM, Mintun MA, Joshi AD, et al. Amyloid deposition, hypometabolism, and longitudinal cognitive decline. *Ann Neurol*. Oct 2012;72(4):578-86. doi:10.1002/ana.23650
16. Landau SM, Lu M, Joshi AD, et al. Comparing positron emission tomography imaging and cerebrospinal fluid measurements of β-amyloid. *Ann Neurol*. Dec 2013;74(6):826-36. doi:10.1002/ana.23908
17. Ge X, Qiao Y, Choi J, et al. Enhanced Association of Tau Pathology and Cognitive Impairment in Mild Cognitive Impairment Subjects with Behavior Symptoms. *J Alzheimers Dis*. 2022;87(2):557-568. doi:10.3233/JAD-215555
18. Aksman LM, Oxtoby NP, Scelsi MA, et al. A data-driven study of Alzheimer's disease related amyloid and tau pathology progression. *Brain*. Dec 01 2023;146(12):4935-4948. doi:10.1093/brain/awad232
19. Dumurgier J, Sabia S, Zetterberg H, et al. A Pragmatic, Data-Driven Method to Determine Cutoffs for CSF Biomarkers of Alzheimer Disease Based on Validation Against PET Imaging. *Neurology*. Aug 16 2022;99(7):e669-e678. doi:10.1212/WNL.0000000000200735
